# SARS-CoV-2 infectivity by viral load, S gene variants and demographic factors and the utility of lateral flow devices to prevent transmission

**DOI:** 10.1101/2021.03.31.21254687

**Authors:** Lennard YW Lee, Stefan Rozmanowski, Matthew Pang, Andre Charlett, Charlotte Anderson, Gareth J Hughes, Matthew Barnard, Leon Peto, Richard Vipond, Alex Sienkiewicz, Susan Hopkins, John Bell, Derrick W Crook, Nick Gent, A Sarah Walker, Tim EA Peto, David W Eyre

## Abstract

**Background:** How SARS-CoV-2 infectivity varies with viral load is incompletely understood. Whether rapid point-of-care antigen lateral flow devices (LFDs) detect most potential transmission sources despite imperfect sensitivity is unknown.

**Methods:** We combined SARS-CoV-2 testing and contact tracing data from England between 01-September-2020 and 28-February-2021. We used multivariable logistic regression to investigate relationships between PCR-confirmed infection in contacts of community-diagnosed cases and index case viral load, S gene target failure (proxy for B.1.1.7 infection), demographics, SARS-CoV-2 incidence, social deprivation, and contact event type. We used LFD performance to simulate the proportion of cases with a PCR-positive contact expected to be detected using one of four LFDs.

**Results:** 231,498/2,474,066 (9%) contacts of 1,064,004 index cases tested PCR-positive. PCR-positive results in contacts independently increased with higher case viral loads (lower Ct values) e.g., 11.7%(95%CI 11.5-12.0%) at Ct=15 and 4.5%(4.4-4.6%) at Ct=30. B.1.1.7 infection increased PCR-positive results by ∼50%, (e.g. 1.55-fold, 95%CI 1.49-1.61, at Ct=20). PCR-positive results were most common in household contacts (at Ct=20.1, 8.7%[95%CI 8.6-8.9%]), followed by household visitors (7.1%[6.8-7.3%]), contacts at events/activities (5.2%[4.9-5.4%]), work/education (4.6%[4.4-4.8%]), and least common after outdoor contact (2.9%[2.3-3.8%]). Contacts of children were the least likely to test positive, particularly following contact outdoors or at work/education. The most and least sensitive LFDs would detect 89.5%(89.4-89.6%) and 83.0%(82.8-83.1%) of cases with PCR-positive contacts respectively.

**Conclusions:** SARS-CoV-2 infectivity varies by case viral load, contact event type, and age. Those with high viral loads are the most infectious. B.1.1.7 increased transmission by ∼50%. The best performing LFDs detect most infectious cases.

**Key points:** In 2,474,066 contacts of 1,064,004 SARS-CoV-2 cases, PCR-positive tests in contacts increased with higher index case viral loads, the B.1.1.7 variant and household contact. Children were less infectious. Lateral flow devices can detect 83.0-89.5% of infections leading to onward transmission.

## Introduction

The global health impact of SARS-CoV-2 is profound.^1^ There is widespread on-going transmission despite control efforts predominantly focused on quarantining symptomatic cases and population-level self-isolation.^2^ The emergence of potentially more transmissible variants, such as B.1.1.7^3^ which has spread widely in the UK, has hampered control. However, vaccine roll-out offers the prospect of reduced disease and transmission.^4^

Intermittent national and regional social distancing and self-isolation measures have been imposed in many countries.^5,6^ Additional self-isolation measures for “contacts” (individuals exposed to SARS-CoV-2) vary by country, but generally last 7-14 days.^7^ While reducing transmission, quarantine/isolation measures have indirectly had many wider effects on economic productivity, well-being^8^ and non-COVID-19-related excess deaths.^9–11^ Not all exposure to SARS-CoV-2 leads to infection, e.g., in some settings only 5-7% of exposed “contacts” develop COVID-19 infection^12,13^ and modelling suggests ∼15% of individuals are responsible for most SARS-CoV-2 transmission.^14^ Therefore, using isolation selectively for those who are most infectious could lessen some of its collateral impacts.^12,13^

Our understanding of how individual infectiousness varies is limited. Several assays for infectivity have been proposed. Functional assays include animal and cell culture models, whereas viral sub-genomic mRNA is a nucleic acid-based measure of infectivity.^15^ Detection of viral protein, i.e. antigen, as assessed by lateral flow devices (LFDs), has been shown to be more closely linked to viral culture infectivity than PCR measurements.^16^ However, few of these surrogate measures of infectivity have been convincingly demonstrated to predict the real-world likelihood of a SARS-CoV-2 infected individual infecting someone else.

Here we use data from the England’s national contact tracing and testing programs to explore the relationship between infectivity and SARS-CoV-2 viral load, as measured by PCR cycle threshold (Ct) values. We identify demographic factors associated with infectivity and assess the impact of the emergence of the B.1.1.7 variant. We apply our results to a population of PCR-positive individuals to estimate the proportion of infectious individuals detected by viral antigen LFDs under a range of performance conditions.

## Methods

Data from community and hospital PCR testing in England between 01-September-2020 and 28-February-2021 were obtained and linked with national contact tracing data by the UK Government Department of Health and Social Care. Data extracts were de-identified prior to analysis and included for PCR-confirmed cases and their contacts: demographic details (age, sex, ethnicity), if symptoms were present for cases and the timing of testing relative to symptom onset, and test results, as well as details on the nature of the contact events.

### Index cases and contacts

We defined index cases as SARS-CoV-2 PCR-positive individuals with a community-based test performed by three high-throughput national testing facilities (“Lighthouse Laboratories” in Milton Keynes, Alderley Park or Glasgow), which reported Ct values indicating viral load. Samples were processed using the same RNA extraction and Thermo Fisher TaqPath PCR platform in each laboratory (targeting S and N genes, and ORF1ab; details in Supplement). Only the first positive result per person was included. Index cases without available Ct values were excluded. The B.1.1.7 variant contains a deletion in the S gene, resulting in S gene target failure (SGTF). Sequencing of SGTF samples showed 89.5% were due to B.1.1.7 by mid-January 2021,^17^ so SGTF was used as a proxy for B.1.1.7.

Contacts of index cases were defined as all individuals notified to the national contact tracing service from the day of the index cases’ positive test until 10 days later with whom the index case had been in close proximity from 48 hours before their symptom onset to 10 days afterwards (further definitions in Supplement). Contacts could be tested PCR-positive through any community or hospital-based test as these were nationally reported.

### Statistical analysis

We aimed to determine factors associated with PCR-positive results in contacts, including the demographics, viral load and SGTF status of the index case. To identify outcomes most likely representing onward transmission from the index case rather than a third party, we excluded contacts named by more than one index case. We also restricted to positive test results obtained 1-10 days following the index case’s test date, i.e., the period when the index case may have been infectious, to exclude earlier results in contacts and avoid contacts who were the source for the index case’s infection. Given these restrictions, the absolute proportion of contacts testing PCR-positive cannot be interpreted as a secondary attack rate, because some onward transmission events are excluded. Where contacts had more than one PCR test within the follow-up window, all were considered to identify positive results.

We used multivariable logistic regression to investigate associations between PCR-confirmed infection in contacts (including contacts whether or not they had PCR tests) and the index case’s Ct value and SGTF status (B.1.1.7 proxy), the contact event nature, the case’s demographics, and incidence and social deprivation index at the contact’s home location. We did not adjust for symptoms in the case, as these may be mediators of the effect of viral load on onward transmission. We used splines to account for non-linearity in continuous variables and screened for all pairwise interactions between main effects (details in Supplement).

We performed sensitivity analyses to test our restriction to contacts tested 1-10 days after each index case and including only contacts with PCR tests. We used unadjusted linear regression to investigate the proportion of the variation in Ct values in contacts that could be explained by the case’s Ct value.

### Simulations of the number of cases identified by antigen LFDs

We used our findings to estimate the proportion of potential transmission events where the source case would have been detected using an antigen LFD, using existing data on the sensitivity of four LFDs: Innova, Deep Blue, Orient gene and Abbott.^18^ For each source case we simulated a positive or negative LFD result by randomly drawing from the probability of a LFD being positive by the source case’s Ct value (see Supplement, Figure S1). Each simulation was repeated 1000 times. Additionally, we ran simulations for a range of hypothetical LFD performances.

### Ethics

The study was conducted as part of national COVID-19 surveillance under the provisions of Section 251 of the NHS Act 2006 and therefore did not require individual patient consent. It was approved by Public Health England (PHE), the UK COVID-19 LFD oversight group and NHS Test and Trace. The protocol for this work was reviewed by the PHE Research Ethics and Governance Group, which is the PHE Research Ethics Committee, and was found to be fully compliant with all regulatory requirements. As no regulatory or ethical issues were identified, it was agreed that a full ethical review would not be needed, and the protocol was approved.

## Results

### Cases and contacts

3,577,246 SARS-CoV-2 PCR-positive results were available from England between 01-September-2020 and 28-February-2021. Of these 1,818,456 (51%) tests were performed by three national laboratories providing high-throughput community testing with a standardised PCR assay, yielding a first positive test per person in 1,796,139 individuals. 27,893 (2%) were excluded as no Ct value was available, 439,482 (24%) had no recorded contacts, and 264,760 (15%) had only recorded contacts shared with other index cases, leaving 1,064,004 index cases in the analysis (Figure 1, Table S1). 487,653 (46%) cases had SGTF, consistent with B.1.1.7, increasing to near 100% by 28-February-2021 (Figure S2).

**Figure 1.**
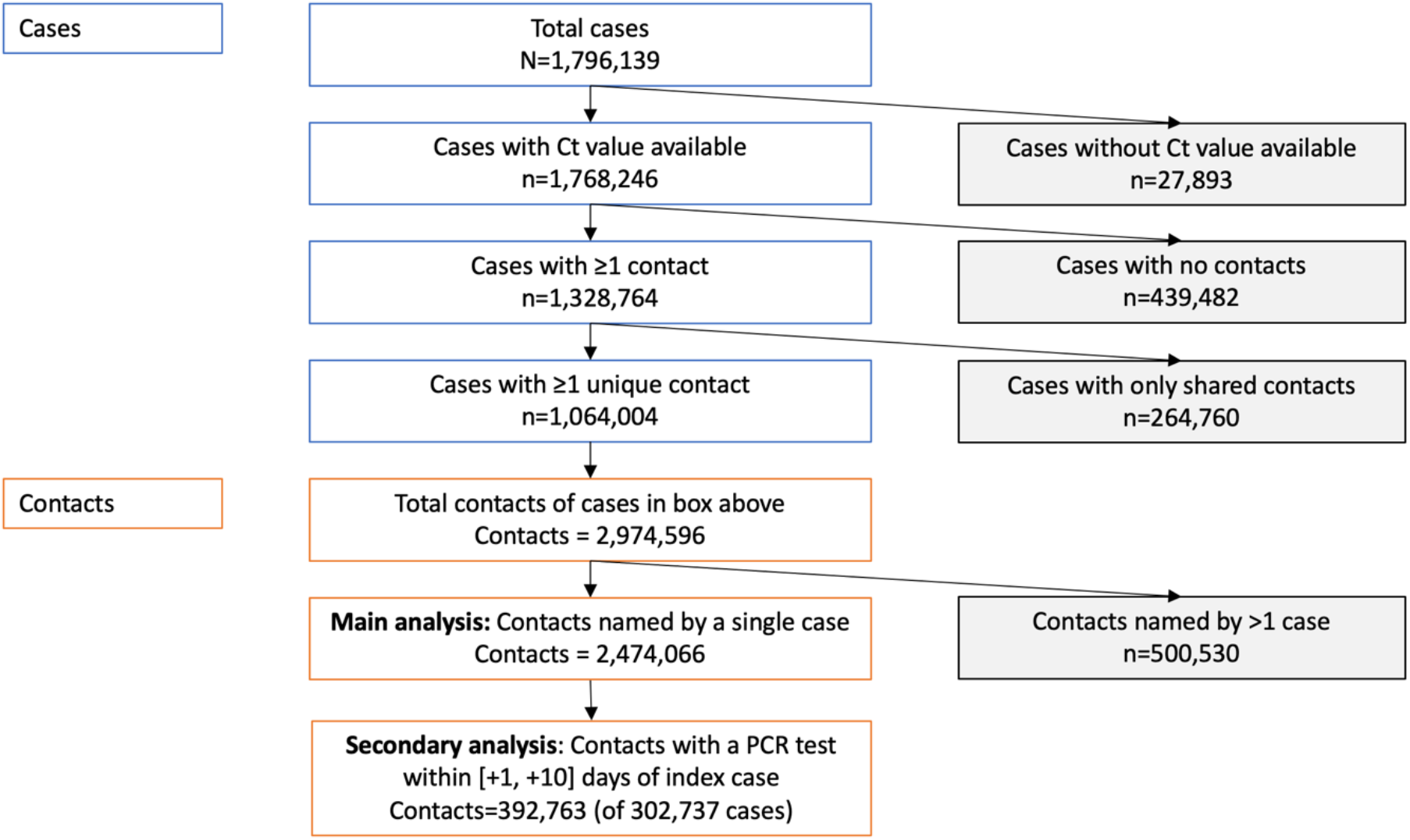
Index cases and contacts in England, 01 September 2020 to 28 February 2021.

The 1,064,004 index cases had 2,974,596 contacts identified within 10 days of their test of whom 918,758 (31%) had a PCR test within ±10 days of the index case and 638,456 (21%) tested PCR-positive. 2,474,066 (83%) contacts were named only by a single case and are included in the analysis; 231,498 (9%) tested PCR positive 1-10 days after the index case’s PCR-positive result, our main outcome measure, i.e., consistent with possible transmission from the index case to the contact.

The median (IQR) age of cases and contacts was 36 (24-51) and 31 (16-49) years respectively, and 54% and 52% with available data were female (Table 1). Most contact events occurred within households (77.4%), followed by visits to households (8.3%), workplaces or education (8.0%), attending events or activities (5.3%), and outdoors (0.3%).

**Table 1.** Demographics and characteristics of the study population. ^1^Frequency (%) or median (IQR).

| Variable | Case,<br>n = 1,064,004<sup>1</sup> | Contact: not PCR-positive within 1-10<br>days,<br>n = 2,242,569<sup>1</sup> | Contact: PCR-positive<br>within 1-10 days,<br>n = 231,497<sup>1</sup> |
| --- | --- | --- | --- |
| Sex |  |  |  |
| Female | 560,557 (53%) | 820,203 (37%) | 114,837 (50%) |
| Male | 476,967 (45%) | 765,538 (34%) | 99,539 (43%) |
| Not specified | 26,480 (2.5%) | 656,828 (29%) | 17,121 (7.4%) |
| Age | 36 (24 - 51) | 30 (15 - 48) | 37 (23 - 52) |
| Not available | 9 | 720,544 | 17,853 |
| Ethnic group |  |  |  |
| Asian | 128,218 (12%) | 77,932 (3.5%) | 9,491 (4.1%) |
| Black | 27,658 (2.6%) | 17,167 (0.8%) | 1,874 (0.8%) |
| Mixed | 27,263 (2.6%) | 19,342 (0.9%) | 2,297 (1.0%) |
| Other | 15,682 (1.5%) | 9,667 (0.4%) | 1,170 (0.5%) |
| White | 728,265 (68%) | 585,255 (26%) | 78,363 (34%) |
| Not available | 136,918 (13%) | 1,533,206 (68%) | 138,302 (60%) |
| Incidence at home address, per<br>100,000 population | 355 (215 - 546) | 348 (207 - 524) | 375 (226 - 581) |
| Not available | 4,124 | 6,444 | 493 |
| Deprivation index at home<br>address (lower = more deprived,<br>of 32,844 areas) | 14,465 (11,374 -<br>18,704) | 14,465 (11,304 -<br>18,649) | 14,593 (11,744 -<br>19,165) |
| Not available | 4,124 | 6,444 | 493 |
| Case symptomatic | 969,942 (91%) |  |  |
| Days from symptom onset to<br>test in case where symptomatic | 2 (1 - 3) |  |  |
| Contact type |  |  |  |
| Events/activities |  | 137,805 (6.1%) | 8,919 (3.9%) |
| Household |  | 1,718,674 (77%) | 196,508 (85%) |
| Household visitor |  | 189,637 (8.5%) | 16,426 (7.1%) |
| Outdoors |  | 8,002 (0.4%) | 317 (0.1%) |
| Work/education |  | 188,451 (8.4%) | 9,327 (4.0%) |
| Days from case diagnosis to<br>contact notification |  | 2 (2- 3) | 2 (1- 3) |
| Days from index case test to<br>contact's test where tested |  | 2 (1 - 4) | 3 (2 - 5) |

### Predictors of PCR-positive results in contacts

On univariable analysis (Table 2, Figures S3-S6), PCR-positive tests in contacts were associated with lower case Ct values (i.e. higher viral loads), SGTF in the index case, higher incidence in the local population, less social deprivation, white ethnicity and male sex. Household contacts were most likely to be PCR-positive. PCR-positive results were least frequent in contacts of children, with highest rates in contacts of older adults.

**Table 2.**
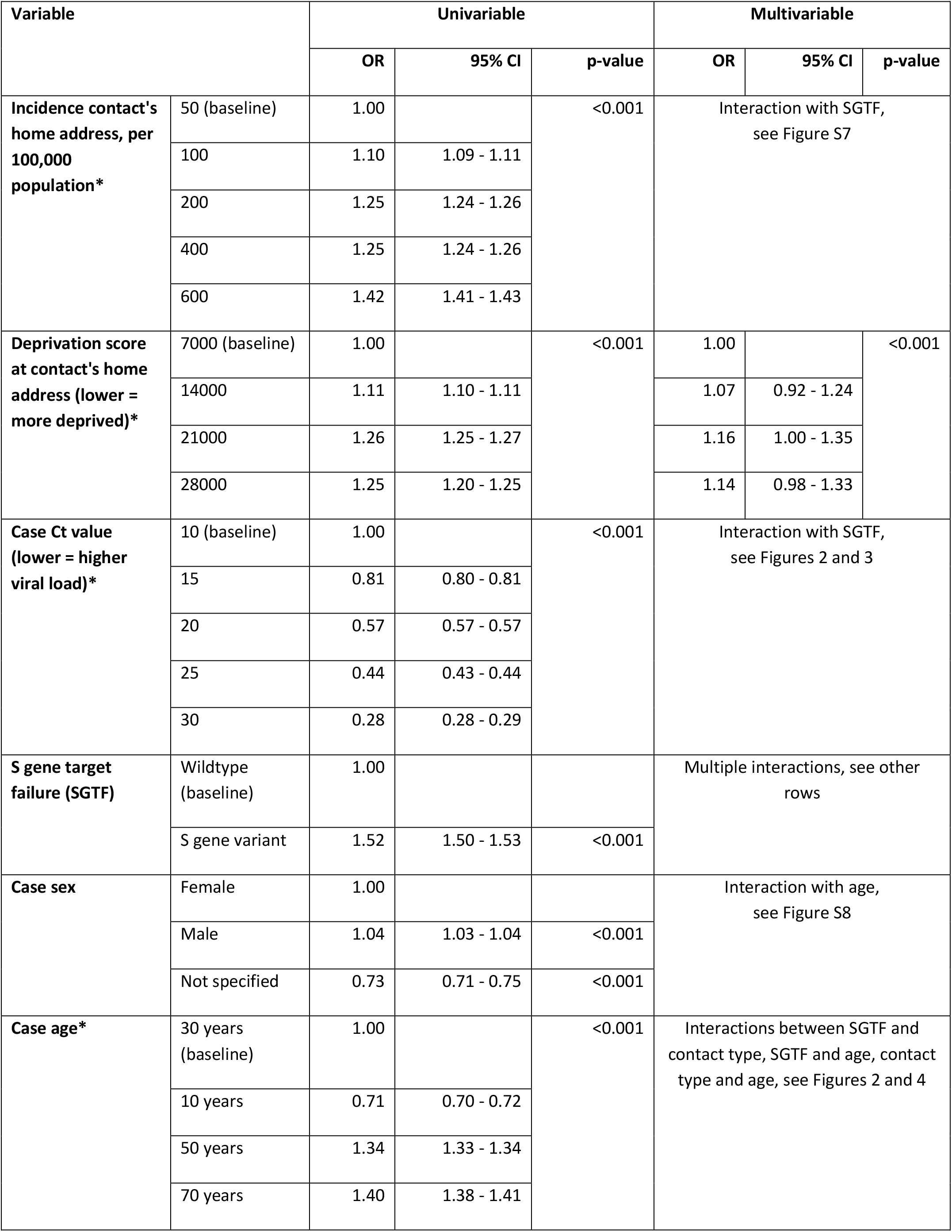
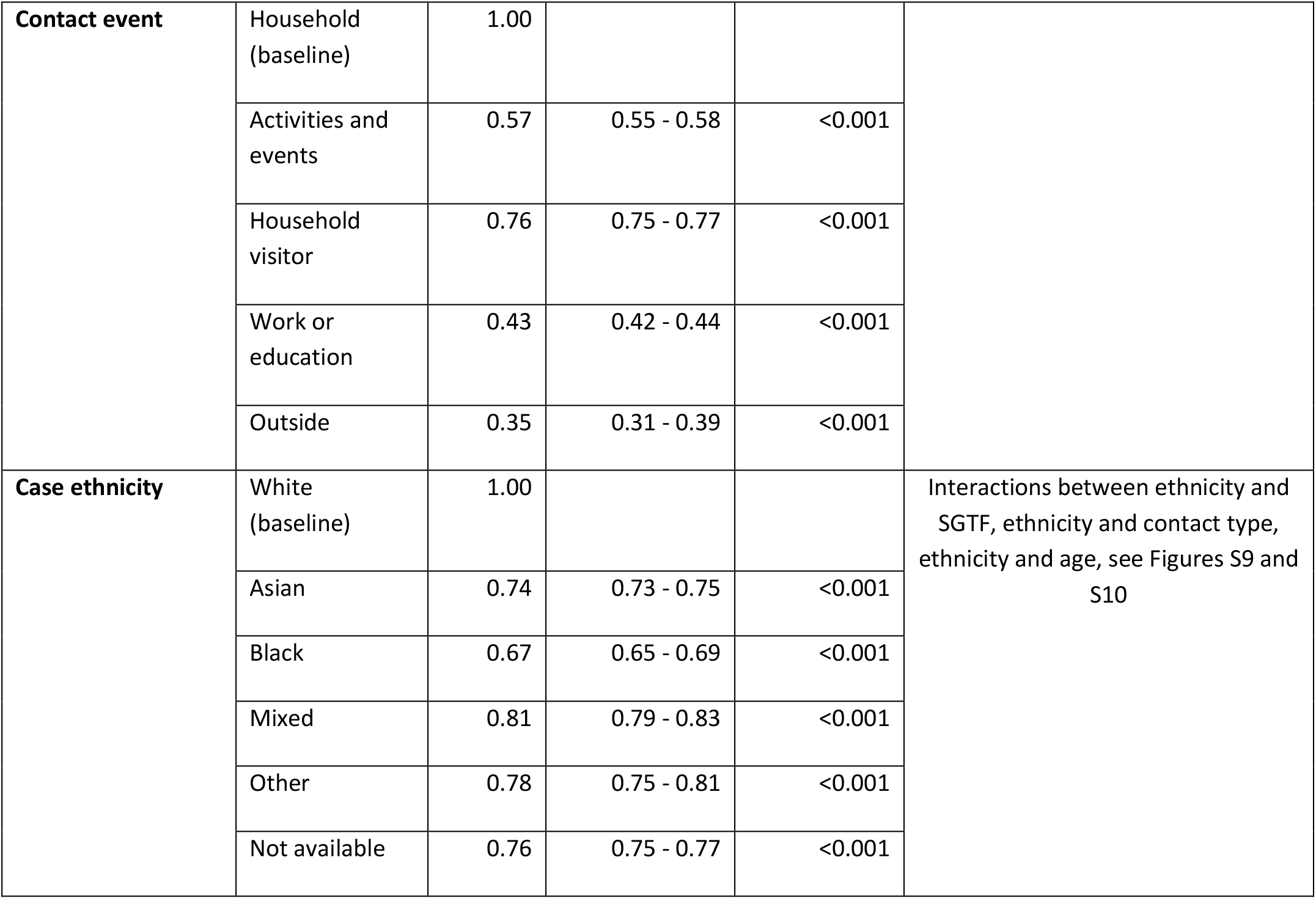
Univariable and multivariable associations with the proportion of contacts testing PCR positive. The lower rates of PCR-positivity seen in cases without a documented sex possibly reflect incomplete contact tracing or poor data quality preventing appropriate linkage of these cases. *Incidence, deprivation score, index case Ct value and case age are all fitted as non-linear effects with 5 default-spaced knots, example values are shown, and univariable relationships plotted in Figures S3-S6. Multivariable results are presented with continuous variables set to their median value and categorical variables set to baseline, figures illustrating relationships with interactions are listed. See Figure S11 for the multivariable relationship for deprivation score.

| Variable |  | Univariable |  |  | Multivariable |  |  |
| --- | --- | --- | --- | --- | --- | --- | --- |
|  |  | OR | 95% CI | p-value | OR | 95% CI | p-value |
| Incidence contact's home address, per 100,000 population* | 50 (baseline) | 1.00 |  | <0.001 | Interaction with SGTF, see Figure S7 |  |  |
|  | 100 | 1.10 | 1.09 - 1.11 |  |  |  |  |
|  | 200 | 1.25 | 1.24 - 1.26 |  |  |  |  |
|  | 400 | 1.25 | 1.24 - 1.26 |  |  |  |  |
|  | 600 | 1.42 | 1.41 - 1.43 |  |  |  |  |
| Deprivation score at contact's home address (lower = more deprived)* | 7000 (baseline) | 1.00 |  | <0.001 | 1.00 |  | <0.001 |
|  | 14000 | 1.11 | 1.10 - 1.11 |  | 1.07 | 0.92 - 1.24 |  |
|  | 21000 | 1.26 | 1.25 - 1.27 |  | 1.16 | 1.00 - 1.35 |  |
|  | 28000 | 1.25 | 1.20 - 1.25 |  | 1.14 | 0.98 - 1.33 |  |
| Case Ct value (lower = higher viral load)* | 10 (baseline) | 1.00 |  | <0.001 | Interaction with SGTF, see Figures 2 and 3 |  |  |
|  | 15 | 0.81 | 0.80 - 0.81 |  |  |  |  |
|  | 20 | 0.57 | 0.57 - 0.57 |  |  |  |  |
|  | 25 | 0.44 | 0.43 - 0.44 |  |  |  |  |
|  | 30 | 0.28 | 0.28 - 0.29 |  |  |  |  |
| S gene target failure (SGTF) | Wildtype (baseline) | 1.00 |  |  | Multiple interactions, see other rows |  |  |
|  | S gene variant | 1.52 | 1.50 - 1.53 | <0.001 |  |  |  |
| Case sex | Female | 1.00 |  |  | Interaction with age, see Figure S8 |  |  |
|  | Male | 1.04 | 1.03 - 1.04 | <0.001 |  |  |  |
|  | Not specified | 0.73 | 0.71 - 0.75 | <0.001 |  |  |  |
| Case age* | 30 years (baseline) | 1.00 |  | <0.001 | Interactions between SGTF and contact type, SGTF and age, contact type and age, see Figures 2 and 4 |  |  |
|  | 10 years | 0.71 | 0.70 - 0.72 |  |  |  |  |
|  | 50 years | 1.34 | 1.33 - 1.34 |  |  |  |  |
|  | 70 years | 1.40 | 1.38 - 1.41 |  |  |  |  |

**Table 2. Univariable and multivariable associations with the proportion of contacts testing PCR** **positive.**
|  |  |  |  |  |  |
| --- | --- | --- | --- | --- | --- |
| **Contact event** | Household (baseline) | 1.00 |  |  |  |
|  | Activities and events | 0.57 | 0.55 - 0.58 | <0.001 |  |
|  | Household visitor | 0.76 | 0.75 - 0.77 | <0.001 |  |
|  | Work or education | 0.43 | 0.42 - 0.44 | <0.001 |  |
|  | Outside | 0.35 | 0.31 - 0.39 | <0.001 |  |
| **Case ethnicity** | White (baseline) | 1.00 |  |  | Interactions between ethnicity and SGTF, ethnicity and contact type, ethnicity and age, see Figures S9 and S10 |
|  | Asian | 0.74 | 0.73 - 0.75 | <0.001 |  |
|  | Black | 0.67 | 0.65 - 0.69 | <0.001 |  |
|  | Mixed | 0.81 | 0.79 - 0.83 | <0.001 |  |
|  | Other | 0.78 | 0.75 - 0.81 | <0.001 |  |
|  | Not available | 0.76 | 0.75 - 0.77 | <0.001 |  |

Adjusted multivariable analysis showed strong evidence of effect modification (interactions) and non-linear relationships, such that associations are best described graphically (Figures 2-3, S7-S11). Index case Ct value was an important determinant of PCR-positive results in contacts, with an approximately linear decline in positive results as Ct value increased, that was independent of the nature of the contact event (Figure 2). For example, amongst household contacts, with other variables set to median values/baseline categories, rates of PCR-positive tests were 11.7% (95%CI 11.5-12.0%) for index case Ct=15 and 4.5% (4.4-4.6%) for Ct=30. Contacts were most likely to test PCR-positive after household contact (percentage of PCR-positive tests, at median Ct value=20.1, 8.7% [95%CI 8.6-8.9%]), followed by visitors to households (7.1% [6.8-7.3%]), contacts at events/activities (5.2% [4.9-5.4%]), then work/education (4.6% [4.4-4.8%]), with outdoor contacts the least likely to test positive (2.9% [2.3-3.8%]).

**Figure 2.**
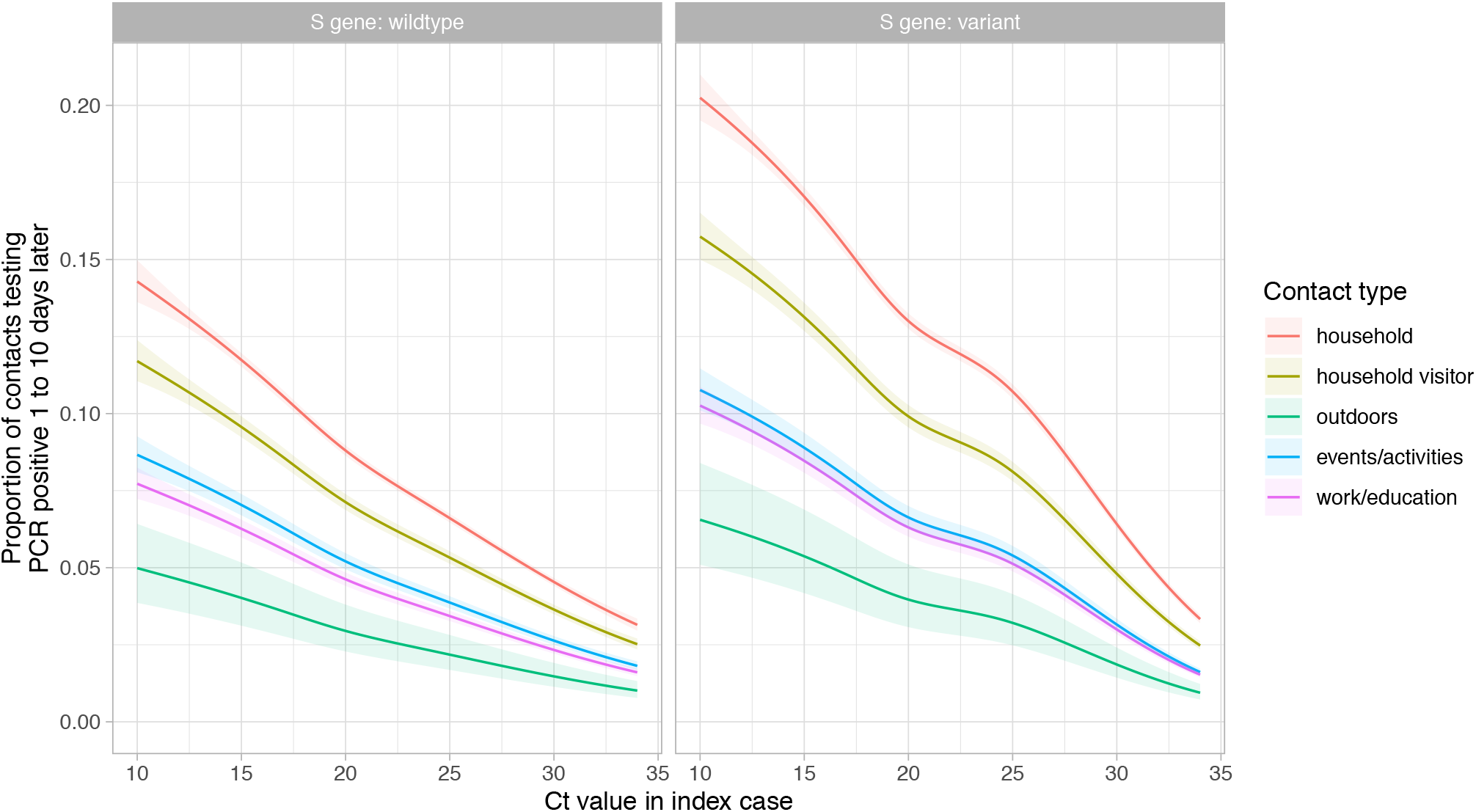
Relationship between PCR cycle threshold (Ct) value in cases and the proportion of their contacts with a PCR positive result, by contact type and S gene target failure. Model predictions are plotted after adjustment for index case age (set to the median value, 35 years), case ethnicity (set to white), index of multiple deprivation score at contact’s home address (set to median, 14,465), incidence at contact’s home address (set to median 350 cases per 100,000 population per week) and index case sex (set to female). The shaded area indicates the 95% confidence interval.

SGTF was associated with increased percentages of contacts testing PCR-positive, by 1.55-fold more (95%CI 1.49-1.60) at index Ct=15, 1.55 (1.49-1.61) at Ct =20 and 1.44 (1.38-1.51) at Ct=30. At Ct values near the upper limit of the assay, the relative increase in PCR-positive results fell to near 1 (Figure 3).

**Figure 3.**
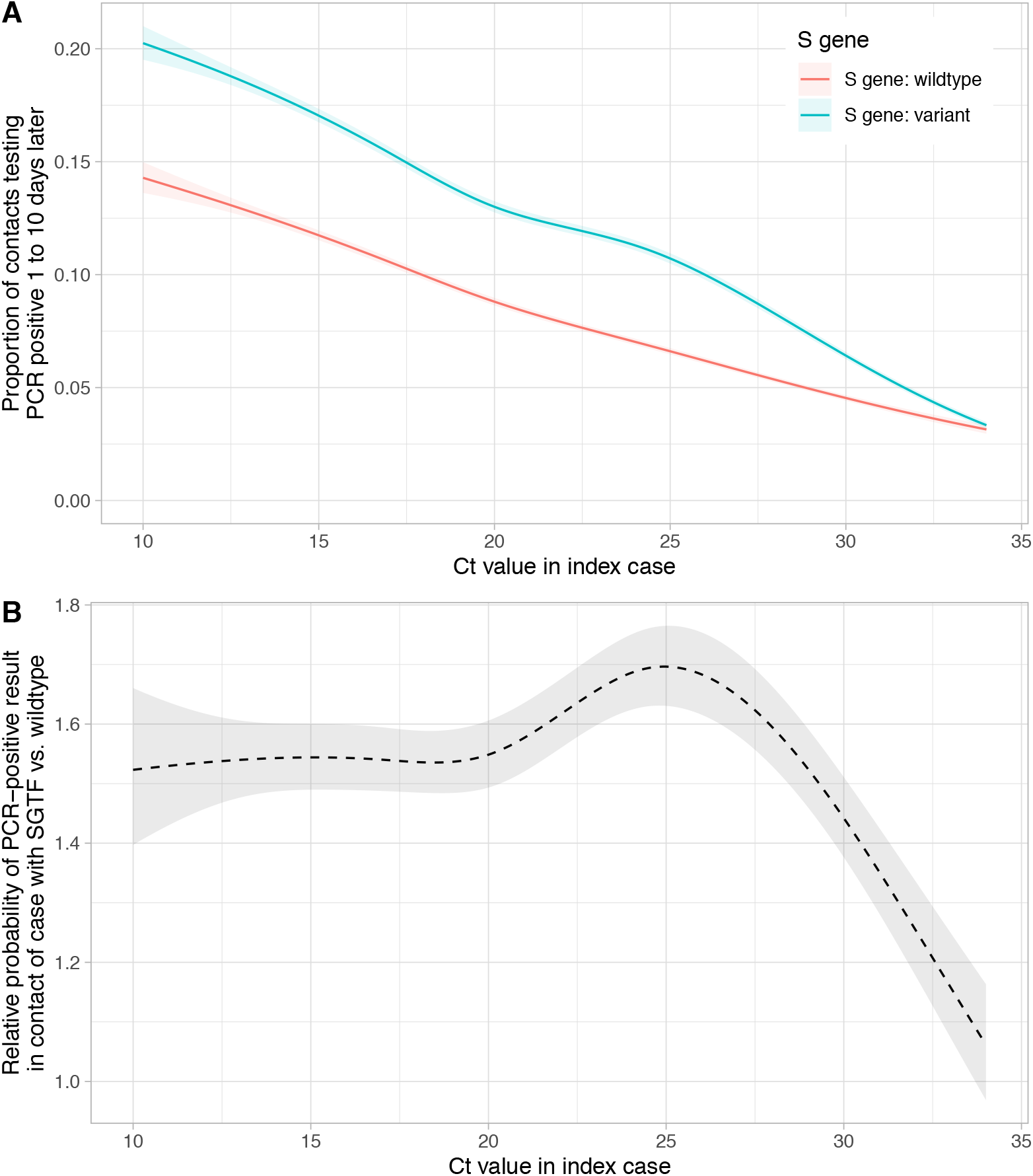
Relationship between PCR-positive results in contacts and index case Ct value and S gene target failure (SGTF) indicative of B.1.1.7 variant. Panel A shows the proportion of contacts testing by PCR-positive. Panel B displays the ratio of the two lines from panel A, i.e., the relative infectiousness of index cases with SGTF vs. without SGTF. Model predictions are adjusted for index case age, sex and ethnicity, contact index of multiple deprivation and incidence as in Figure 2.

Contacts of children were the least likely to test positive, particularly following contact outdoors or at work or in education (Figure 4). Most contact types had similar rates of PCR-positive results across adult ages, except for household contact where risk increased as age increased above 35 years and contact at work/education, events/activities and outdoors where risk of a PCR-positive result was highest in adults in their 20s.

**Figure 4.**
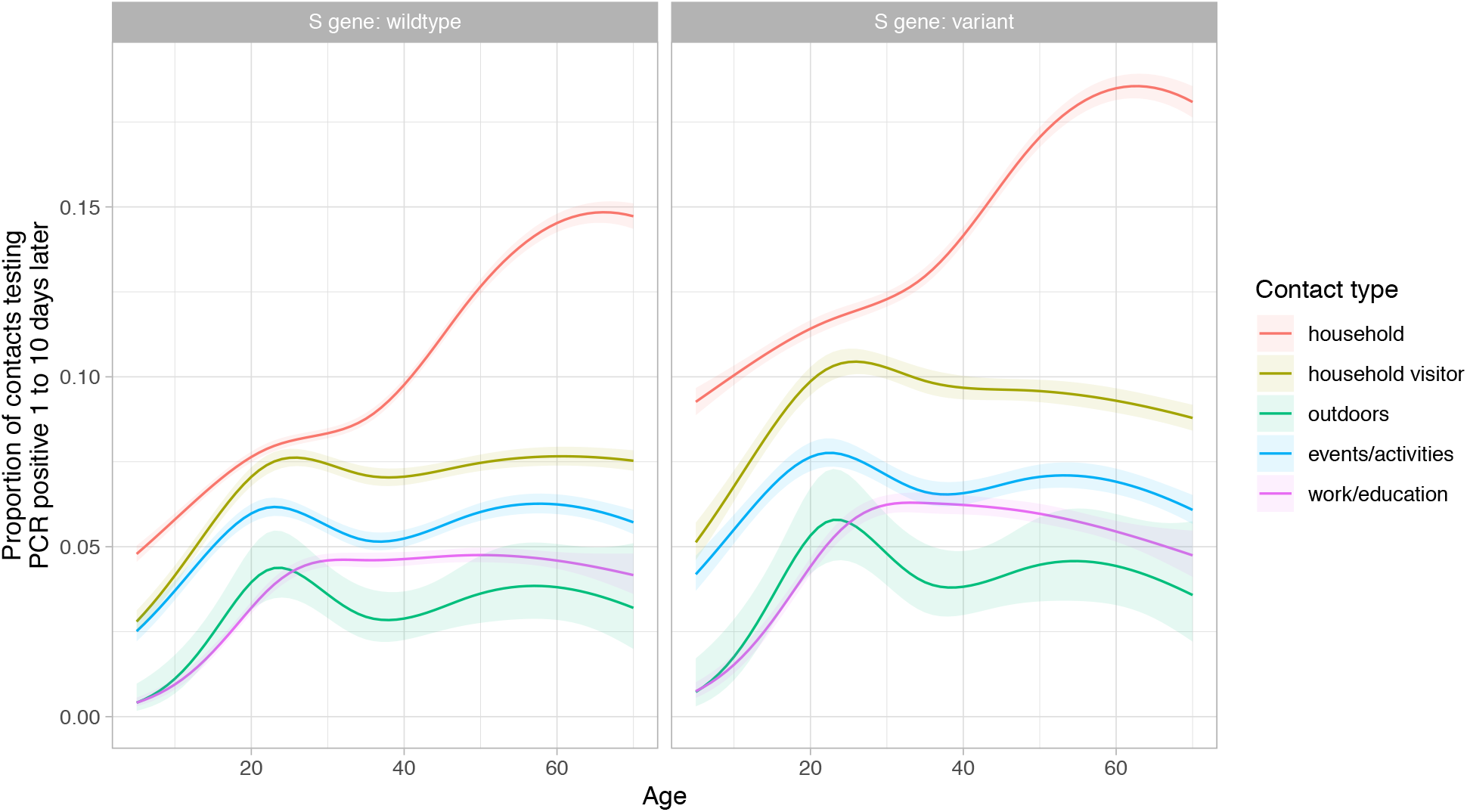
Relationship between index case age and the proportion of their contacts with a PCR positive result, by contact type and S gene target failure. Model predictions are plotted after adjustment for Ct value (set to the median Ct value, 20.1), and other variables as in Figure 2.

Associations between PCR-positive results in contacts and sex varied with age (Figure S8). Broadly, increasing incidence increased PCR-positive contacts, likely reflecting increased acquisition from third parties. There were fewer PCR-positive contacts in areas of greater social deprivation (Figure S11) and amongst Black, Asian and minority ethnic groups (Figure S9-S10).

A sensitivity analysis supported the 1-10 day follow-up window for PCR results in contacts (Figure S12). Case Ct values explained only a small proportion of the variability in contact Ct values (unadjusted linear regression coefficient 0.14 [95%CI 0.13-0.14, p<0.001], R-squared = 0.02).

### Predictors of PCR-positive results in contacts attending PCR testing

In a sensitivity analysis restricted to contacts who had a PCR test (Table S2, Figures S13-S19), similar relationships were seen between PCR-positive results and index case Ct values, contact type and SGTF (Figures S19). While rates of PCR-positive results remained highest in older adult household contacts, there was attenuation of the lower rates seen in children, consistent with main analysis findings of less transmission from children arising from less testing being required or undertaken in contacts of children (Figure S18). In contrast to the main analysis, contacts of all non-white ethnic groups (Table S2) and those living in more deprived areas (Figure S17) were more likely to be PCR-positive, potentially due to differences in access to and use of testing by different ethnic and socioeconomic groups.

### Proportion of cases with PCR-positive contacts detected by LFDs

Overall, 85.4% (197,677/231,497) of case-contact pairs with PCR-positive contacts, i.e., plausible onward transmission, had case viral loads of ≥10,000 RNA copies/ml (i.e. Ct ≤24.4) versus 75.2% of all cases (800,020/1,064,004). Index cases with SGTF had lower Ct values, except for results near the detection threshold (Figure S20).

As antigen LFD sensitivity varies by viral load, we used the distribution of viral loads in case-contact pairs with a PCR-positive contact to simulate the proportion of such cases who would have been detected using antigen LFDs (Figure 5). The Deep Blue LFD would have detected 85.9% (95%CI 85.8-86.0%) of cases who plausibly subsequently transmitted to a contact, the Innova LFD 83.0% (82.8-83.1%), the Orient Gene LFD 89.5% (89.4-89.6%) and the Abbott LFD 85.8% (85.7-86.0%). Performance was very similar before and after B.1.1.7 expansion (Table S3). The performance characteristics required to detect varying proportions of transmission sources by a novel LFD are illustrated in Figure S21.

**Figure 5.**
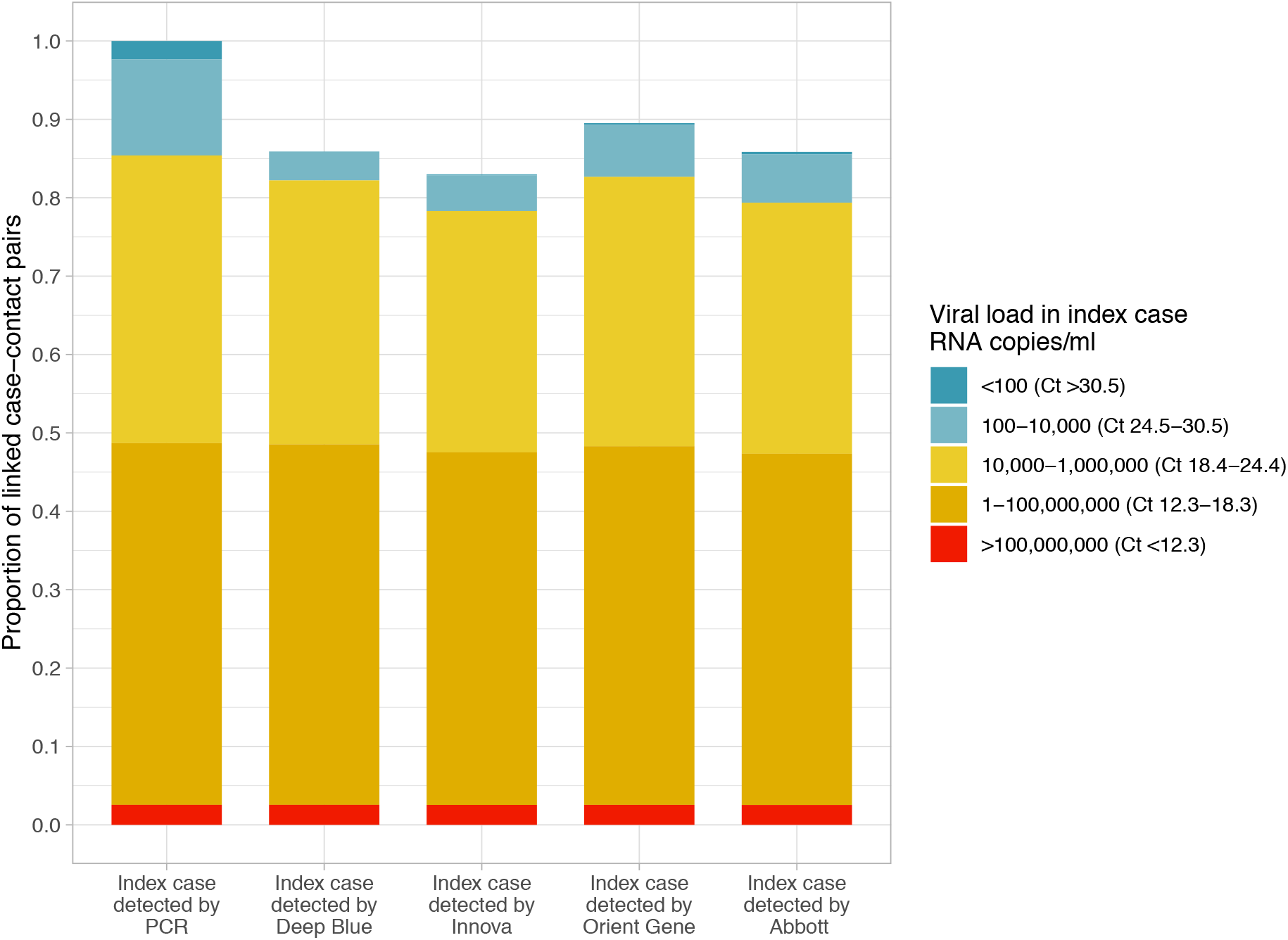
Simulated proportion of cases with a PCR-positive contact detected using four lateral flow devices (LFD). The proportion of cases detected by PCR viral load group is shown in the PCR column. The number of cases with a PCR-positive contact who would be detected using each LFD is shown for 4 LFDs.

## Discussion

We have performed a large-scale analysis of combined SARS-CoV-2 contact tracing and testing data from England involving >2 million contacts of PCR-confirmed cases. We show SARS-CoV-2 infectivity is associated with index case viral load, including after adjustment for demographic factors and type of contact event. SGTF, a proxy for the B.1.1.7 variant, increased transmission by ∼50% at most viral loads. Onward transmission from children was relatively uncommon compared to adults, although this may partly be due to less testing in their contacts. We confirm earlier findings that household contact is associated with greater rates of transmission compared to workplace, educational or recreational contact outside of homes.^19,20^

Except SGTF, it is noteworthy that we found no evidence of significant interactions between Ct values and any other variables in the analysis, i.e. the effect of viral load on infectivity is generalisable across populations and settings. These results are consistent and add to a recent smaller cohort study.^21^

Consistent with other reports^3^ we found that SGTF increased the proportion of contacts testing PCR-positive, by around 55% at high viral loads (Ct values of 10-20), rising with moderate viral loads to a maximum of 75% (Ct=25) before declining again to below 10% at low viral loads (Ct=34). SGTF also affected how the likelihood of transmission varied with age, contact event type and ethnicity. The higher relative infectiousness at moderate viral loads may represent increased infectiousness of individual virions at viral loads where stochasticity is more important compared to higher viral loads. The attenuation of the relative infectiousness at high Ct values partly arises from greater numbers of wildtype strains exhibiting SGTF due to stochastic failure to detect a single gene at low viral loads. As lower viral loads are less infectious, it may also reflect more PCR-positive contacts acquiring infection from third parties, such that the characteristics of the index case matter less. This is supported by the proportion of contacts testing PCR-positive not tending to zero at very low viral loads.

85.4% PCR-positive contacts had an index case with a viral load of ≥10,000 RNA copies/ml (Ct≤24.4). Hence, 85.4% of infections in contacts are potentially attributable to the 75.2% of cases overall with a viral load of ≥10,000 RNA copies/ml. While such data could be used to drive differential interventions to prevent onward transmission with a particular focus on those with high viral loads, our findings suggest that most infected individuals still have some risk of transmitting onwards based on Ct values.

However, we show that several LFDs are sufficiently sensitive to detect most cases that led to onward transmission. These tests offer potential advantages, in returning a result in 15-30 minutes, not requiring laboratory infrastructure and costing significantly less than PCR tests. However post-analytic infrastructure is still needed to collect results. Using the estimated sensitivity of four LFDs, we estimate they would detect 83.0%-89.5% of cases leading to onward transmission. While such performance is not sufficient to replace PCR for testing of all symptomatic individuals, use of LFDs in addition to existing testing, particularly of those who otherwise would not be tested at all (including those without symptoms), would allow many of the most infectious individuals to be identified earlier, potentially preventing onward transmissions and helping to drive reproduction numbers below 1, despite imperfect performance against PCR. The specificity of each LFD is another important consideration, particularly as incidence falls; the false positive rate for the Innova LFD has been previously reported as 0.32% (95%CI 0.20-0.48%),^18^ and large-scale evaluations of the other LFDs are on-going. In settings where the positive predictive value of an LFD is insufficiently high, confirmatory PCR testing may be required.

Our study has important limitations. Firstly, ascertaining infection in contacts depends on the contact being reported by the case and the contact being tested. In the UK, PCR testing is only recommended for those with symptoms and therefore we do not ascertain most asymptomatic infections. Whilst Ct values are generally slightly lower in those without symptoms,^22^ they may nevertheless contribute substantially to transmission.^23^ Additionally, access to testing depends on social and demographic factors, e.g. the relationships between PCR-positive results in contacts and ethnicity varied if we conditioned on contact attendance for a PCR test (Table 2 vs. Table S2).

Secondly, our classification of contact events is relatively simple, e.g., we do not have any direct measures of human behaviour, such as proximity or duration of contact. We also do not account for the dynamic nature of viral loads over time,^24^ relying on a single measurement at varying times post infection. Despite this, the time from symptom onset to testing in the cases was relatively consistent, median (IQR) 2 (1-3) days, such that measured Ct values plausibly represent similar stages of the illness in cases. We use only a single assay to determine Ct values, but have calibrated this to allow comparison with other platforms.

Finally, it was not possible to account for unobserved third-party transmission, although we designed our study population to minimise this risk. This likely means that some contact events identified as possible transmission events may actually not be the source of the infection in the contact. It is likely that proportionally this effect is greatest at lower viral loads (higher Ct values), as the likelihood of transmission rises with viral load.

In summary, we provide strong evidence that SGTF increases SARS-CoV-2 transmission and that SARS-CoV-2 infectivity increases with increasing viral load. We show that the relative strength of the effect of viral load is consistent across ages, ethnicities, and different types of contact events. Despite this association, most individuals have Ct values compatible with onward transmission.^25^ Nevertheless, LFDs can detect most individuals who are potential transmission sources. This supports wider use of LFDs as rapid and regular screens to detect infectiousness in populations at high risk of acquisition, including recent contacts of cases. Further prospective studies will be required to demonstrate whether targeted isolation and/or contact tracing, together with wider use of LFDs in combination with vaccination are effective in preventing ongoing SARS-CoV-2 transmission.

## Data Availability

The data are not publicly available given their scale and confidential nature.

