## Supplement for "SARS-CoV-2 infectivity by viral load, S gene variants and demographic factors and the utility of lateral flow devices to prevent transmission"

### Supplementary methods

#### Infectivity datasets

Community PCR results are submitted by Government testing centres to a central database via the National Pathology exchange (NPEx) and hospital PCR results are submitted by hospitals routinely to Public Health England's national Second Generation Surveillance System (SGSS). Results from NPEx and SGSS were pooled prior to analysis. Contact tracing data were collected by NHS Test and Trace.

The data extract was made on 11 March 2021, such that all cases had ≥10 days of follow up for contacts.

#### Data linkage

Data linkage was undertaken by the UK Government Department of Health and Social Care using patient identifiers. PCR data from NPEX and SGSS were matched Test and Trace data using multi-field exact joins on either [first name, last name, date of birth and postcode] or [phone number, date of birth, postcode].

#### Data definitions

Contacts were defined as follows in line with national guidelines:[1] a person who has been close to someone who has tested PCR-positive for COVID-19 anytime from 2 days before the person was symptomatic up to 10 days from onset of symptoms. The nature of the contact could include:

- Living in the same household OR
- Face to face contact (within 1 metre for any length of time) or skin to skin contact or someone the case coughed on OR
- Within 1 metre for 1 minute of longer OR
- Within 1-2 metres for more than 15 minutes OR
- Sexual contacts OR
- Travel in the same vehicle or a plane

Ethnicity was summarised using 5 ethnic groups defined by the UK government ("white", "mixed or multiple ethnic groups", "Asian or Asian British", "Black, African, Caribbean or Black British", "Other ethnic group").

Local COVID-19 incidence at the time of each contact event was determined using data for the lower tier local authority (LTLA) containing the contact's home address. Incidence was calculated in two week blocks using the number of cases diagnosed per week at that LTLA per 100,000 population.

Contact deprivation indices were sourced from the latest census, averaged for each LTLA.[2]

### Laboratory methods

Ct values were available for community tests undertaken by the UK's Lighthouse Laboratories in Milton Keynes, Alderley Park and Glasgow. PCR testing was undertaken using the same validated Thermo Fisher TaqPath assay at each site (targeting S and N genes, and ORF1ab) following extraction of 200µl of viral transport media on the Thermo Fisher Kingfisher extraction platform, yielding 60µl post-extraction of which 6µl was used in the PCR reaction. All samples were analyzed separately without pooling of samples. Testing is performed according to UK Accreditation Service and ISO 15189 standards.

PCR outputs were analysed using UgenTec FastFinder 3.300.5, with an assay-specific algorithm and decision mechanism that allows conversion of amplification assay raw data from the ABI 7500 Fast into test results with minimal manual intervention. Samples are called positive if at least a single N-gene and/or ORF1ab are detected (although S-gene cycle threshold (Ct) values are determined, S-gene detection alone is not considered sufficient to call a sample positive) and PCR traces exhibit an appropriate morphology.

Mean Ct values were calculated across all non-missing targets.

To enable comparison of data across PCR assays the Qnostics SARS-CoV-2 Analytical Q Panel 01 (Qnostics, Glasgow, UK) was used to calibrate Ct values from the Thermo Fisher assay into equivalent synthetic RNA viral load (VL) in copies per ml. The resulting equation for converting Ct values into viral loads for the Thermo Fisher TaqPath assay was $\log_{10}(VL) = 12.0 - 0.328*Ct$.

### Statistical analysis

We used multivariable logistic regression to investigate the association between PCR-confirmed infection in contacts (including contacts whether or not they attended for PCR testing) and the Ct value in the index case, the presence of SGTF in the index case (as a proxy for B.1.1.7 infection), the nature of the contact, the case's demographics and incidence and social deprivation index at the contact's home address location. We did not adjust for symptoms in the case, as these may be mediators of the effect of viral load on onward transmission.

Only case-contact pairs with complete data were included in the final regression models. To allow for inclusion of categorical variables with missing data an additional category of "Not available" was included. In total 6966/2,474,066 (0.3%) of case-contact pairs were excluded, 31 missing case age, and 6937 missing contact incidence and multiple deprivation index data.

We used backwards elimination based on the Bayesian information criterion (BIC) to select model main effects, using natural cubic splines to account for non-linearity in continuous variables (up to 5 default-spaced knots, choosing the final number of knots based on BIC). We screened for all pairwise interactions between main effects, retaining interactions that minimised BIC. We used robust standard errors clustering by index case to account for some contacts sharing the same source. Univariable results are reported using same number of splines as selected in multivariable model.

Data analysis was performed using R, version 4.0.4.

To test our restriction of contacts PCR results, to those performed between 1 and 10 days of each index case, we also conducted a sensitivity analysis where we fitted a simpler regression model. We included all PCR results for contacts tested between -4 and 14 days of the case's PCR result and included the time difference between tests as a categorical variable in days and a linear term for index case Ct value in the model (as the full model shows an approximately linear relationship with Ct value). By using an interaction term between these two terms we could estimate how the odds ratio associated with a unit change in Ct value changed by days between tests. Given the relationship between Ct values and infectivity, lower odds ratios per unit increase in Ct value provided evidence that the index case was more likely the source for the contact, and allow the plausibility of different time windows between the index case and contact results to be tested.

### LFD sensitivity by viral load

We used previously reported data and estimates of the sensitivity of four LFDs by viral load from a community-based evaluation.[3] Data were available for the Innova SARS-CoV-2 Antigen Rapid Qualitative Test (Innova), Anhui Deepblue Medical Technology COVID-19 (Sars-CoV-2) Antigen Test kit (Colloidal Gold) (Deep Blue), Abbott PanBio COVID-19 Ag (Abbott) and the Zhejiang Orient Gene Biotech Co. Coronavirus Ag Rapid Test Cassette (Orient Gene).

Data were available from 420 Innova, 177 Deep Blue, 99 Abbott and 95 Orient Gene LFD tests performed in individuals diagnosed with SARS-CoV-2 infection by PCR within the last 5 days. Self-obtained combined nasal and oropharyngeal swabs were analysed. Contemporaneous paired swabs were obtained for repeat PCR testing. Repeat PCR testing was undertaken using the Roche Cobas SARS-CoV-2 test and platform. To enable comparisons with Thermo Fisher TaqPath PCR Ct values we used data on 254 additional samples tested using both extraction and PCR assay methods to enable the following conversion to be derived using linear regression: (Cobas Ct) = 5.5 + 1.0*(Thermo Fisher Ct).

As the sensitivity of each LFD varies by the viral load present, it is not possible to accurately summarise performance with a single sensitivity measurement in a way that can generalise. Therefore, for each LFD we fitted a logistic regression model, to generate the probability of a positive LFD test for a given Ct value or viral load. The results of each regression model are shown in Figure S1, which includes an indication of the uncertainty in the estimated sensitivity at each viral load.

We next applied these viral load-sensitivity curves to our contact tracing dataset. We performed a separate set of simulations for each LFD as follows: for each source case we simulated a positive or negative LFD result by randomly drawing from the probability of the LFD being positive by the index case's Ct value. Each simulation was repeated 1000 times and summary results presented.

To account for uncertainty in the performance of each LFD, for each iteration we sampled a different viral load-sensitivity curve from the range of uncertainty shown in Figure S1. This was done by using the parameters of the underlying logistic regression model, with the slope and intercept sampled as a pair using the mean, standard error and covariance of each parameter.

Additionally, we generated simulations for a range of alternative hypothetical lateral flow test performances.

### Supplementary figures

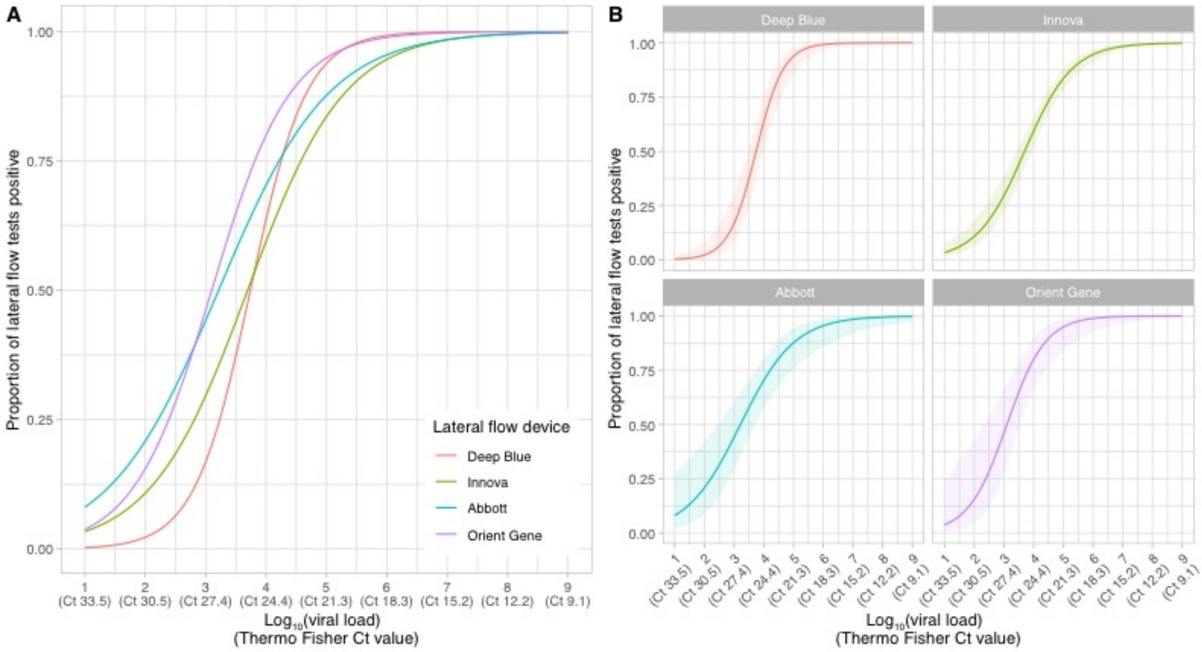


**Figure S1. Sensitivity of four lateral flow devices by viral load**. Thermo Fisher TaqPath assay equivalent Ct units are shown using the formula: $\log_{10}$(viral load) = 12.0 - 0.328*Ct. Panel A shows the fitted relationship, and panel B the 95% confidence intervals for each curve.

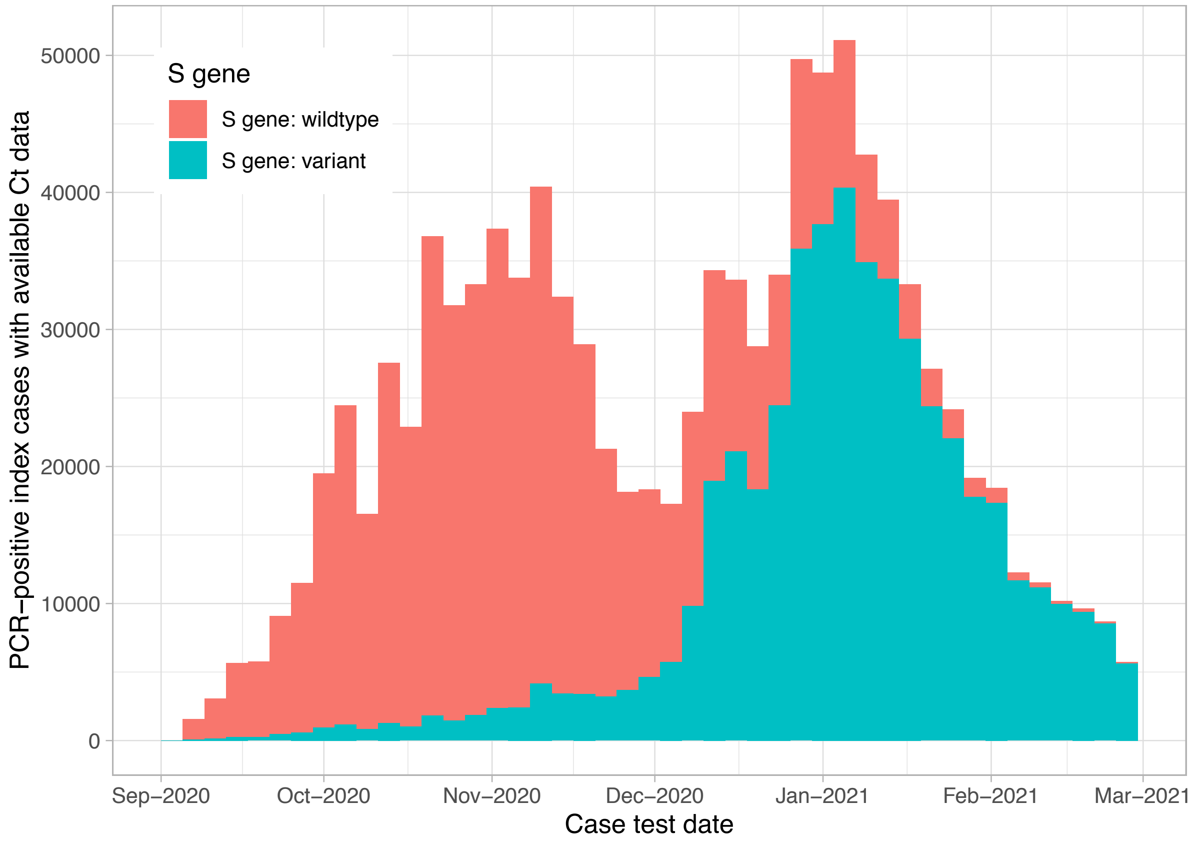


**Figure S2. Incidence of SARS-CoV-2 positive PCR results from England from 3 national laboratories providing community-based testing**. A total of 1,768,246 results are plotted. Results from 27,893 tests without available Ct values are not shown. Each bar represents a 4 day period. Samples with S gene target failure (SGTF), indicative of the B.1.1.7 variant in this setting are shown in blue.

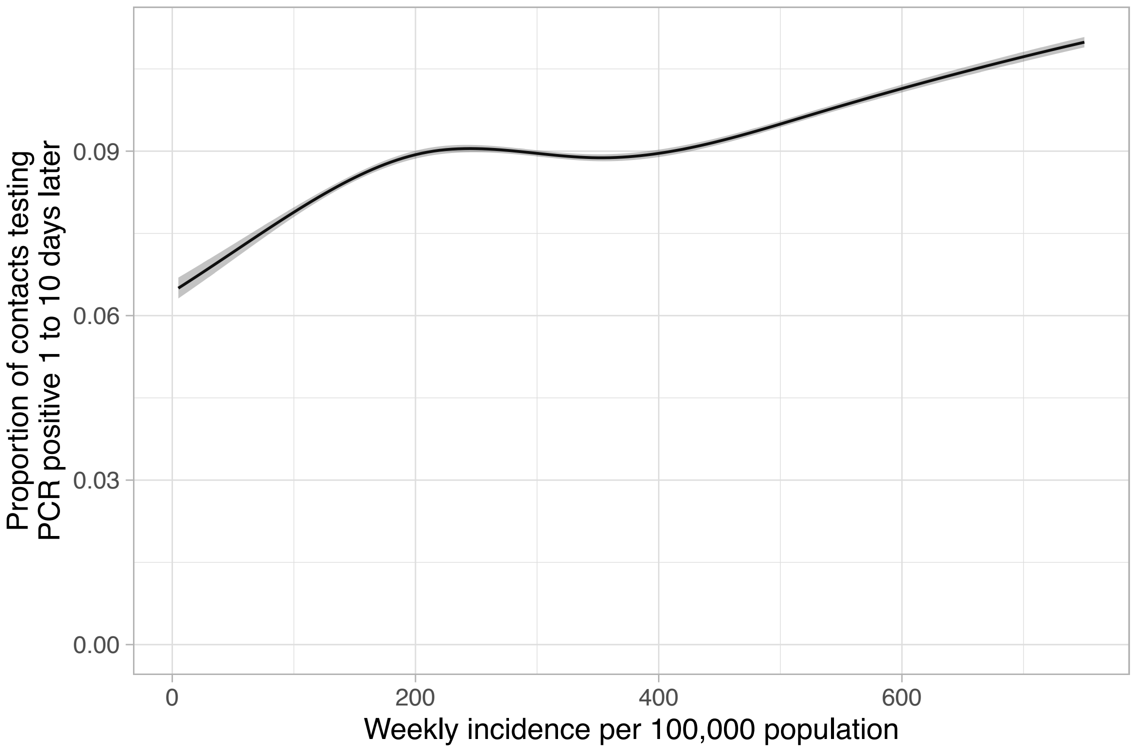


**Figure S3. Univariable relationship between incidence at the contact's home address and proportion of contacts testing PCR positive**. Plotted based on 5-knot spline with default spaced knots, the ribbon shows the 95% confidence interval.

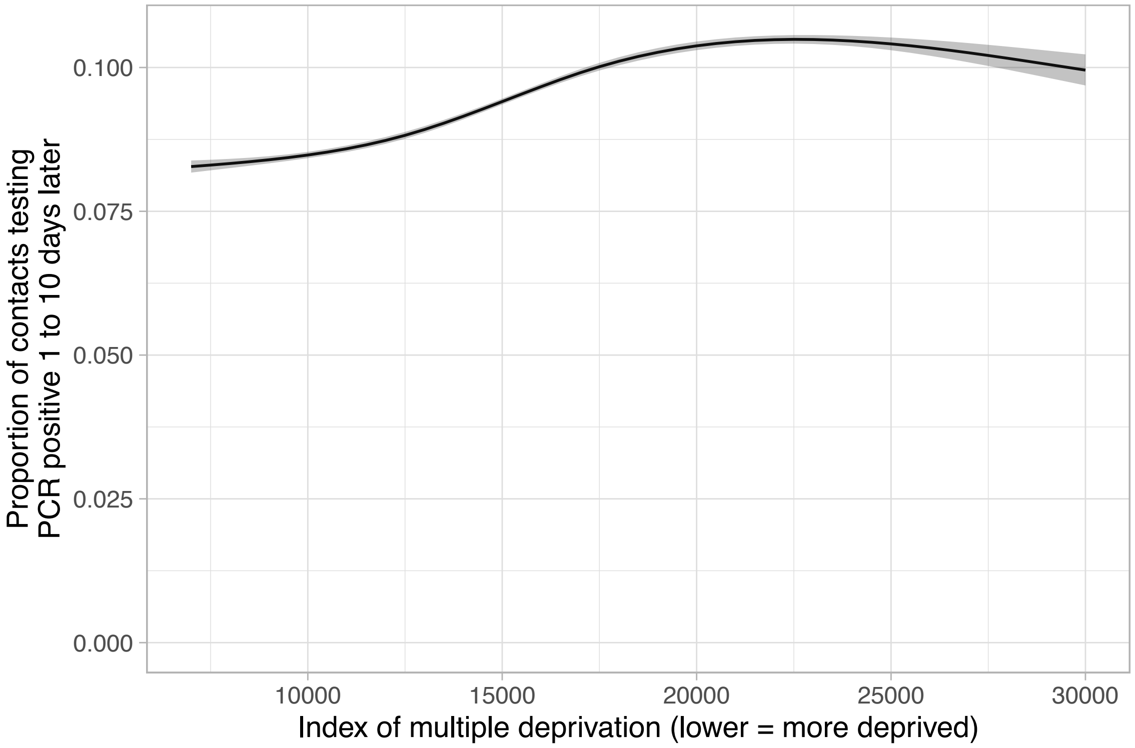


**Figure S4. Univariable relationship between index of multiple deprivation at the contact's home address and proportion of contacts testing PCR positive**. Plotted based on 5-knot spline with default spaced knots, the ribbon shows the 95% confidence interval.

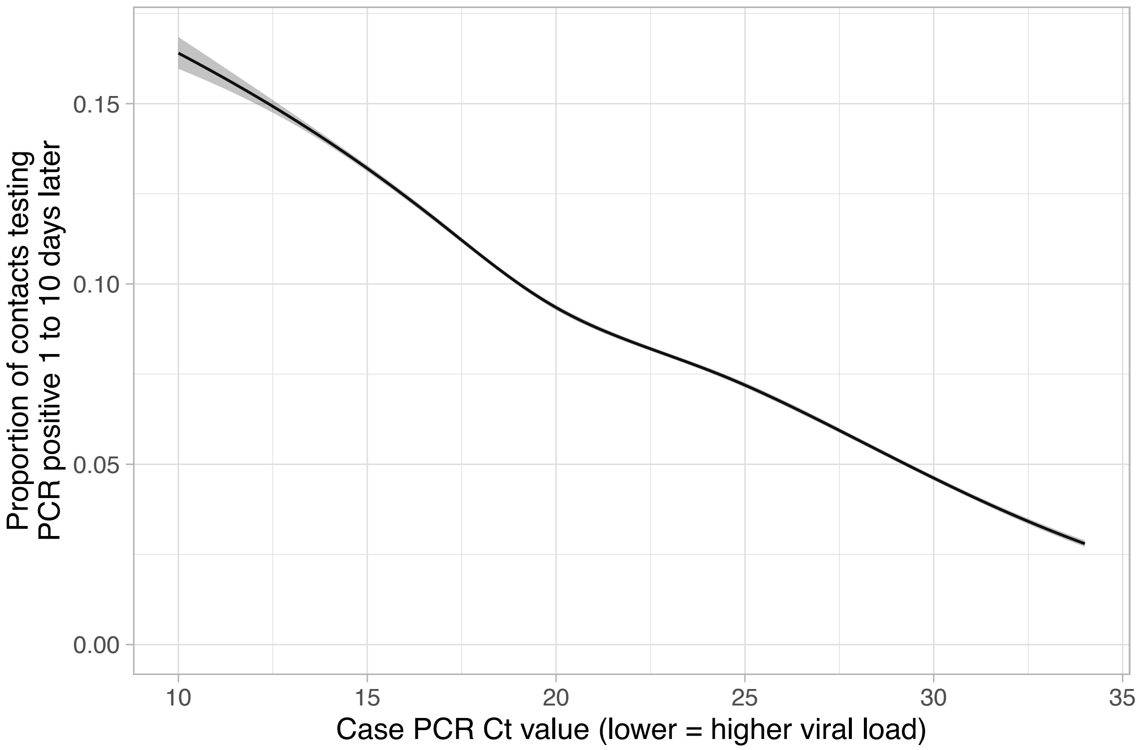


**Figure S5. Univariable relationship between index case Ct value and proportion of contacts testing PCR positive**. Plotted based on 5-knot spline with default spaced knots, the ribbon shows the 95% confidence interval.

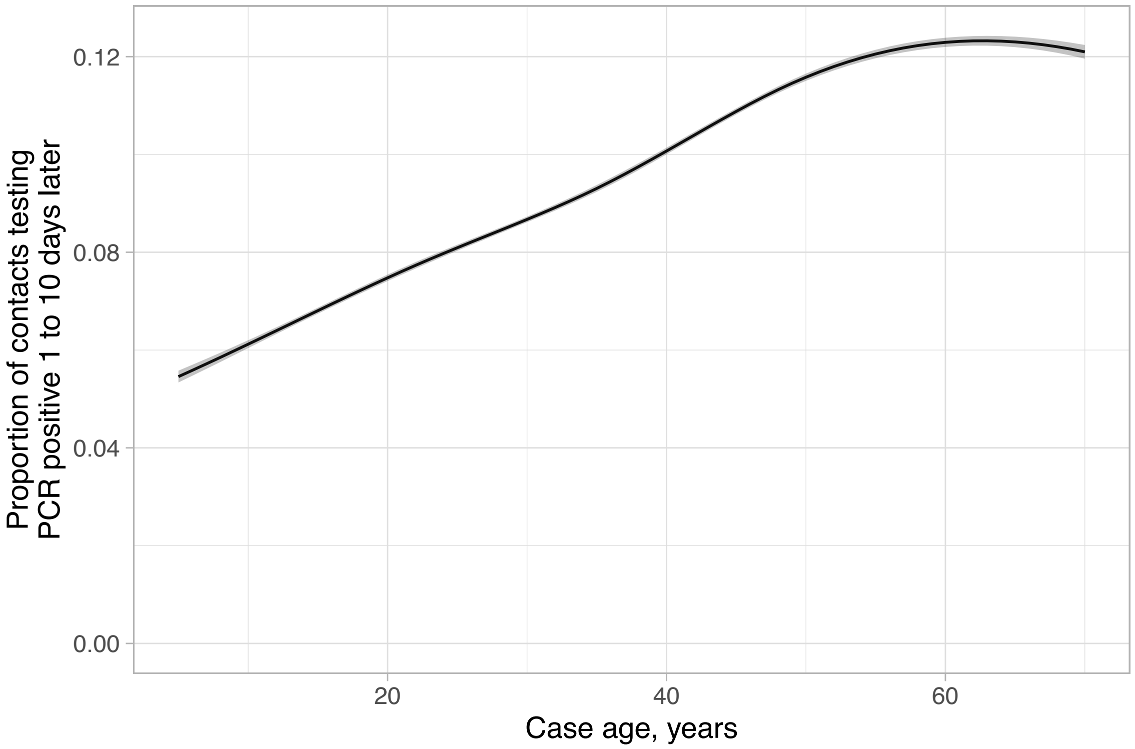


**Figure S6. Univariable relationship between age of the index case and proportion of contacts testing PCR positive**. Plotted based on 4-knot spline with default spaced knots, the ribbon shows the 95% confidence interval.

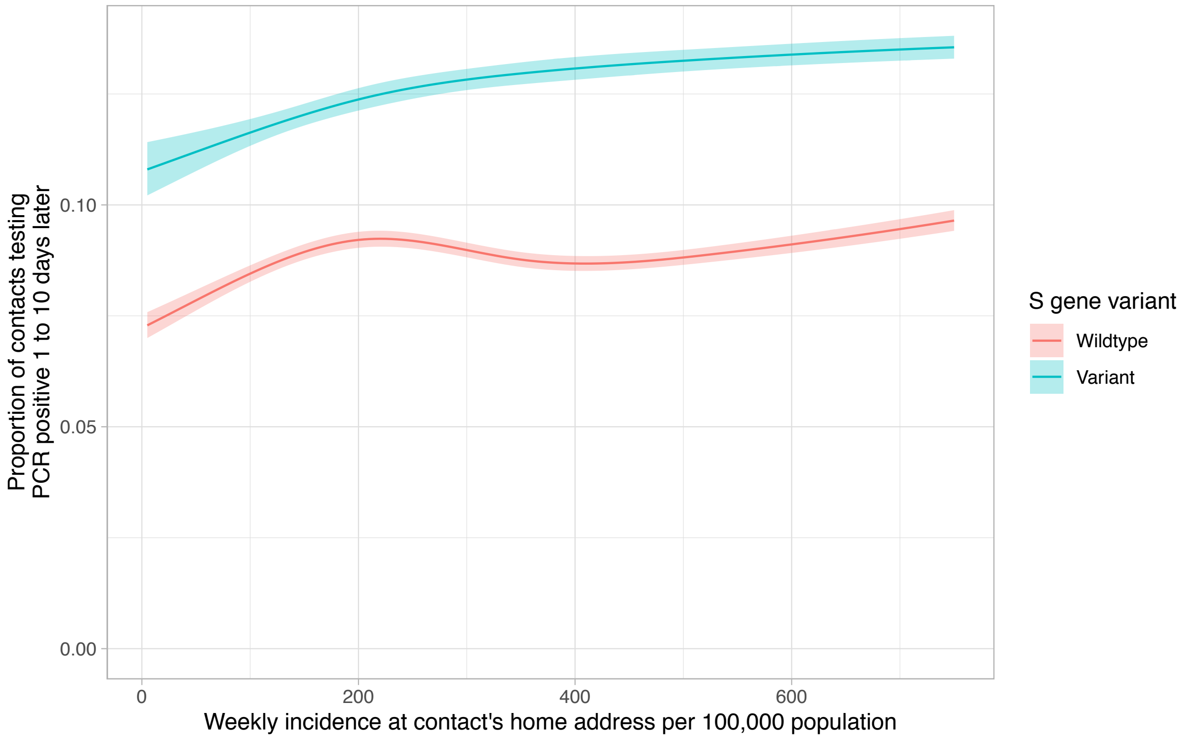


**Figure S7. Relationship between weekly incidence at contact's home address and the proportion of their contacts with a PCR positive result, by S gene target failure**. Model predictions are plotted after adjustment for index case age (set to the median value, 35 years), case ethnicity (set to white), index of multiple deprivation score at contact's home address (set to median, 14,465), Ct value (set to median 20.1) and index case sex (set to female). The shaded area indicates the 95% confidence interval.

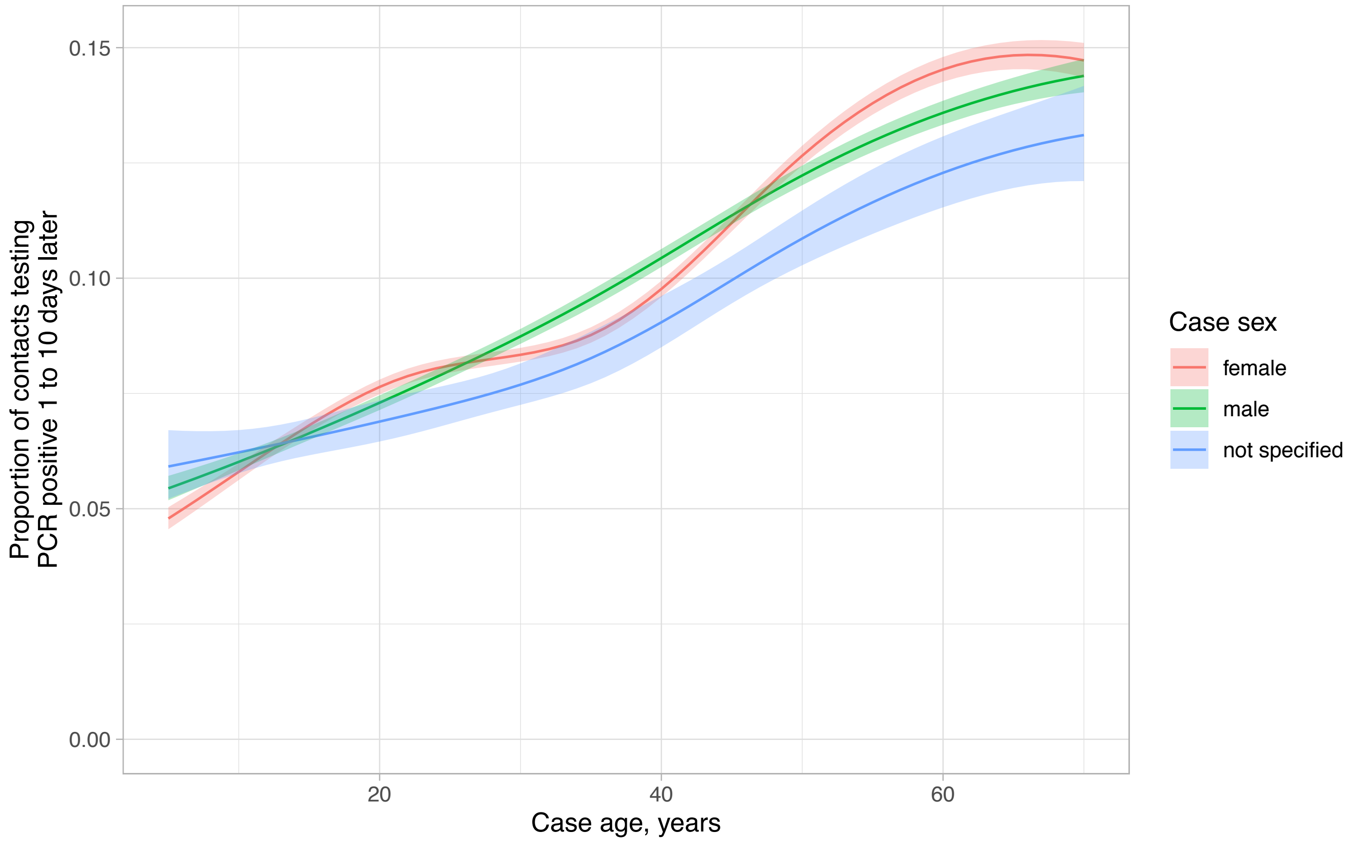


**Figure S8. Relationship between index case age and sex and the proportion of their contacts with a PCR positive result**. Model predictions are plotted after adjustment for case ethnicity (set to white), index of multiple deprivation score at contact's home address (set to median, 14,465), Ct value (set to median 20.1) and index SGTF (set to wildtype). The shaded area indicates the 95% confidence interval.

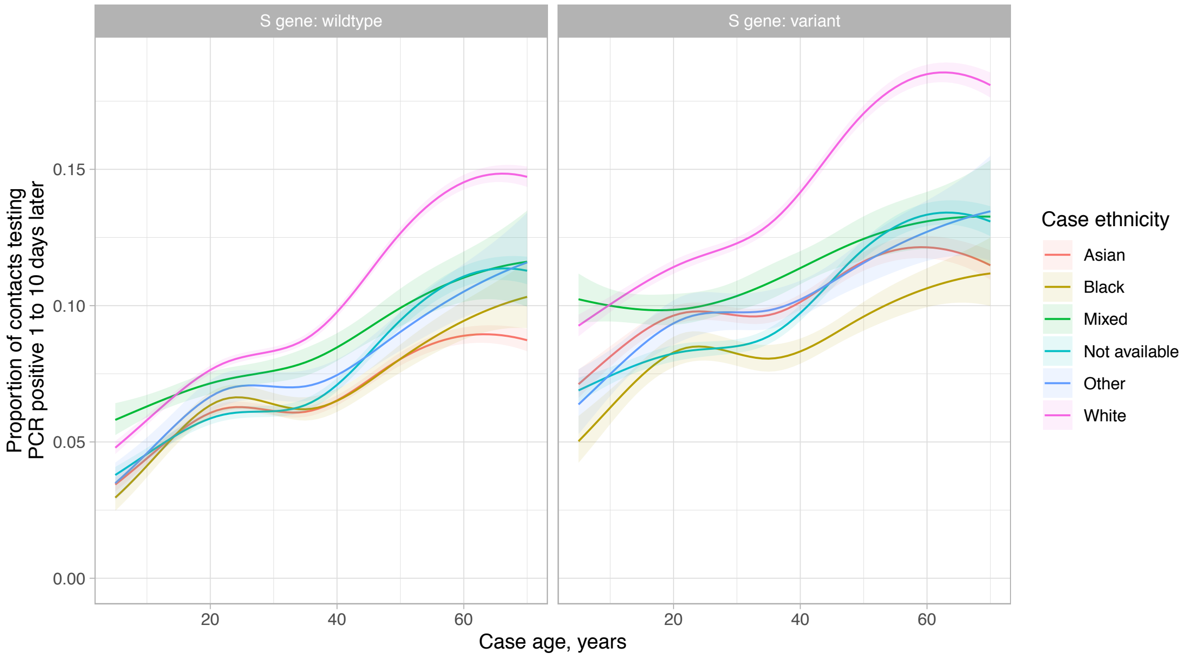


**Figure S9. Relationship between index case age and ethnicity and the proportion of their contacts with a PCR positive result, by SGTF**. Model predictions are plotted after adjustment as above.

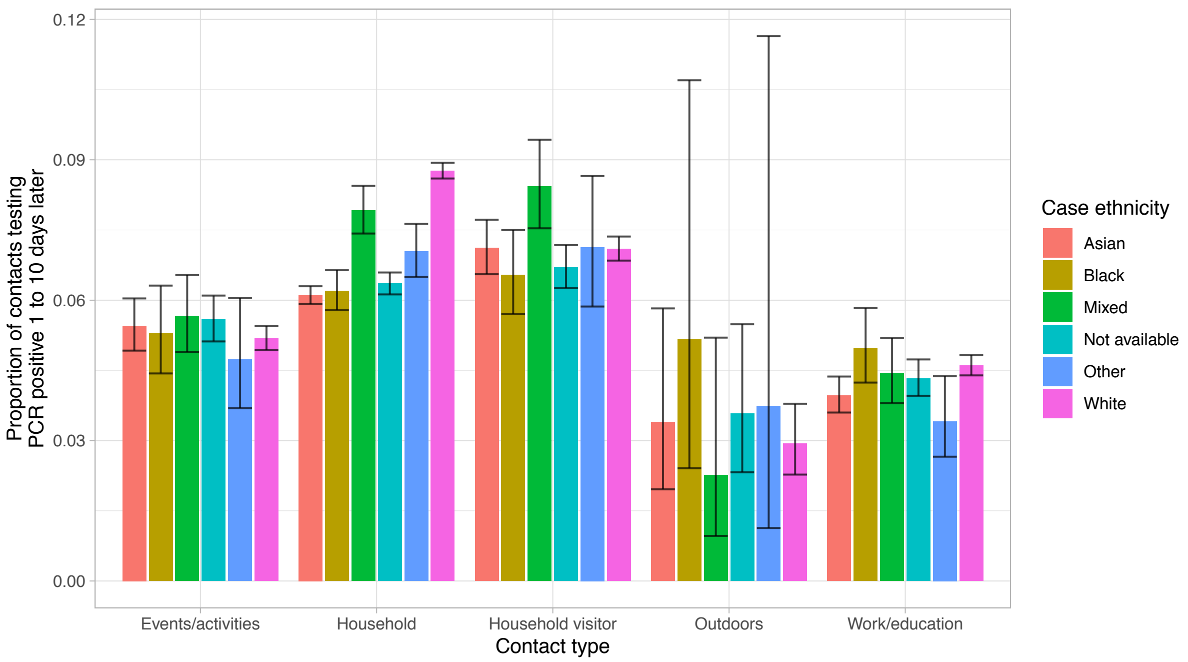


**Figure S10. Relationship between index case ethnicity and the proportion of their contacts with a PCR positive result, by contact type**. Model predictions are plotted after adjustment as above.

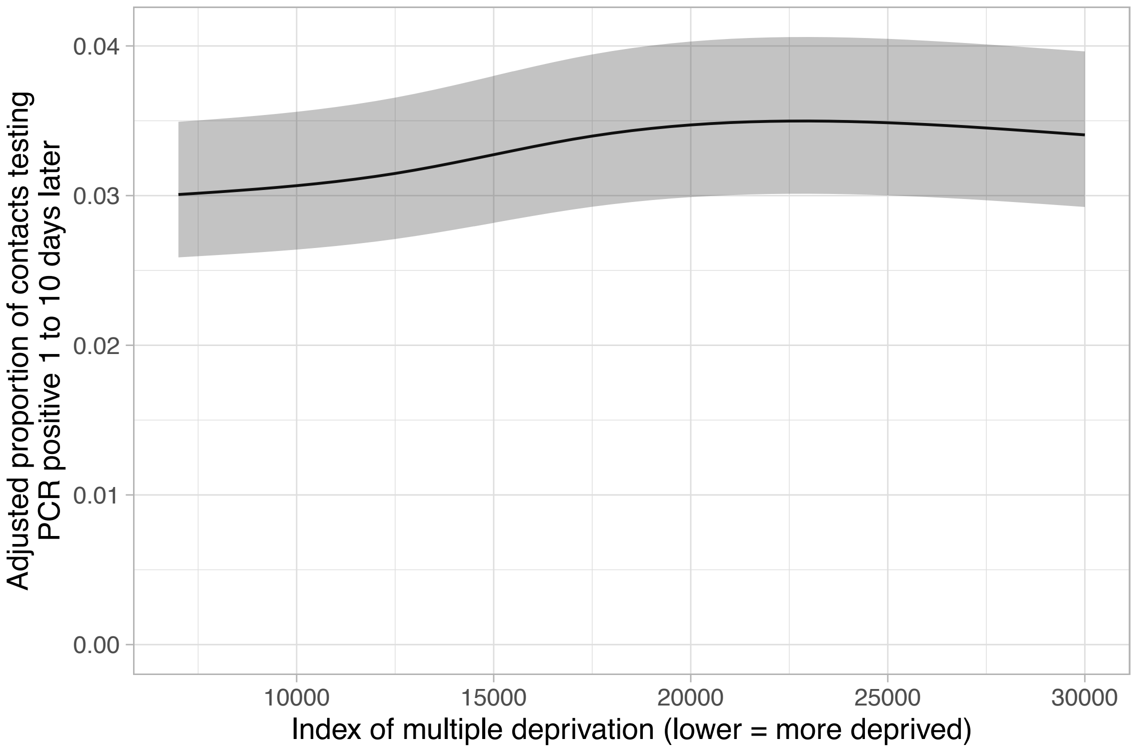


**Figure S11. Multivariable relationship between index of multiple deprivation at contact's home address and the proportion of contacts with a PCR positive result**. Model predictions are plotted after adjustment as above.

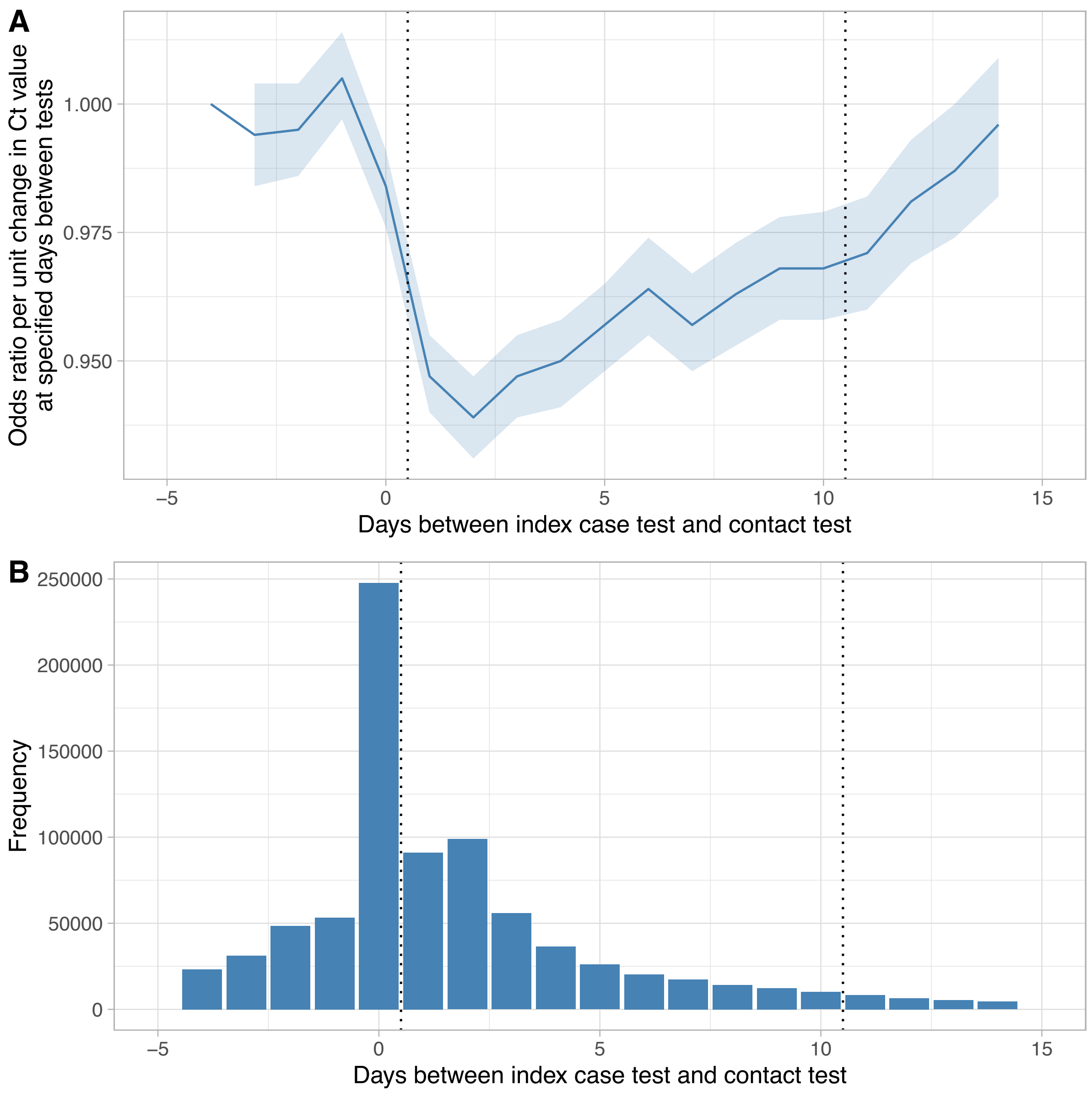


**Figure S12. Sensitivity analysis: odds ratio per unit change in Ct value at specified days between case and contact PCR tests (panel A) and frequency of days between PCR tests (panel B).** Odds ratios in panel A are calculated from a model fitting case Ct value as a linear predictor of positive results in contacts. A categorical term is included for each different number of days between the index case test and contact test from -4 days to +14 days. An interaction is included between the two terms to allow the odds ratio at each distinct number of days to be calculated. The dashed vertical lines indicate the [+1, +10] day window used for the main analysis. Given the relationship between Ct values and infectivity, lower odds ratios per unit increase in Ct value provided evidence that the index case was more likely the source for the contact, with the strongest evidence for the index case as the source for days 1-4 post the index case's test. Although the odds ratio decreased >4 days post index case test, the proportion of contacts tested at longer time intervals was relatively small (panel B), meaning that findings were relatively insensitive to the upper boundary chosen for the follow-up window.

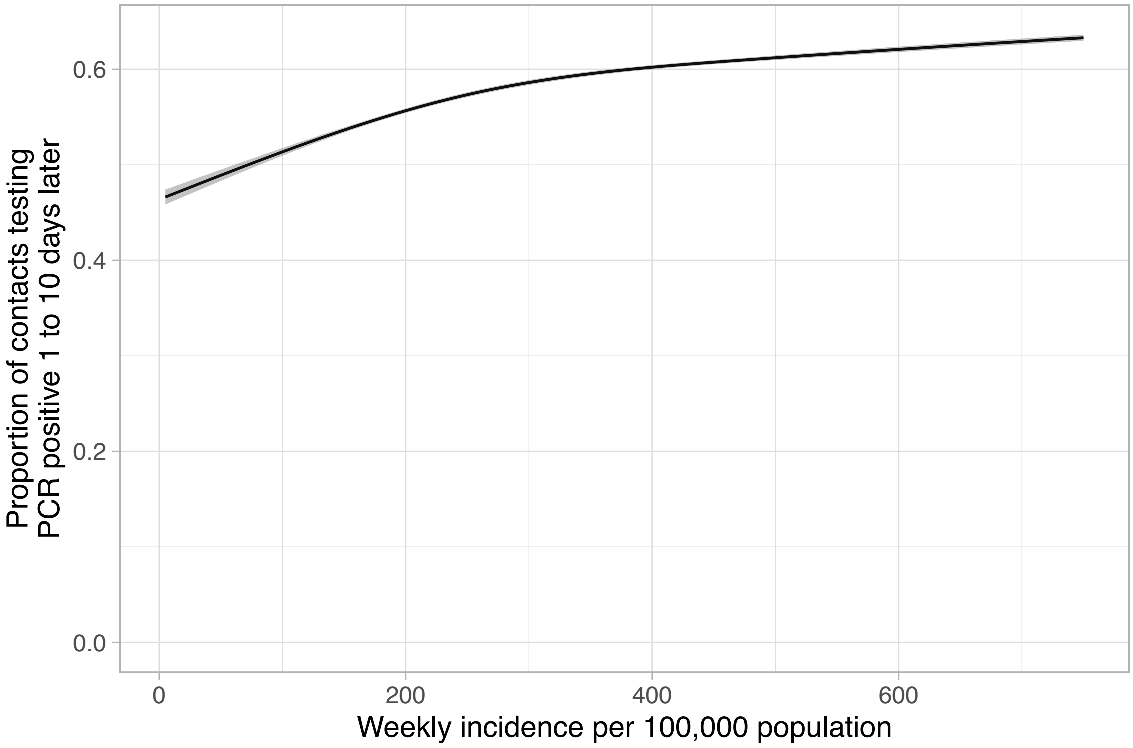


**Figure S13. Sensitivity analysis (restricting to contacts with a PCR test): univariable relationship between weekly incidence at the contact's home address and the proportion of contacts testing PCR positive**. The shaded are shows the 95% confidence interval.

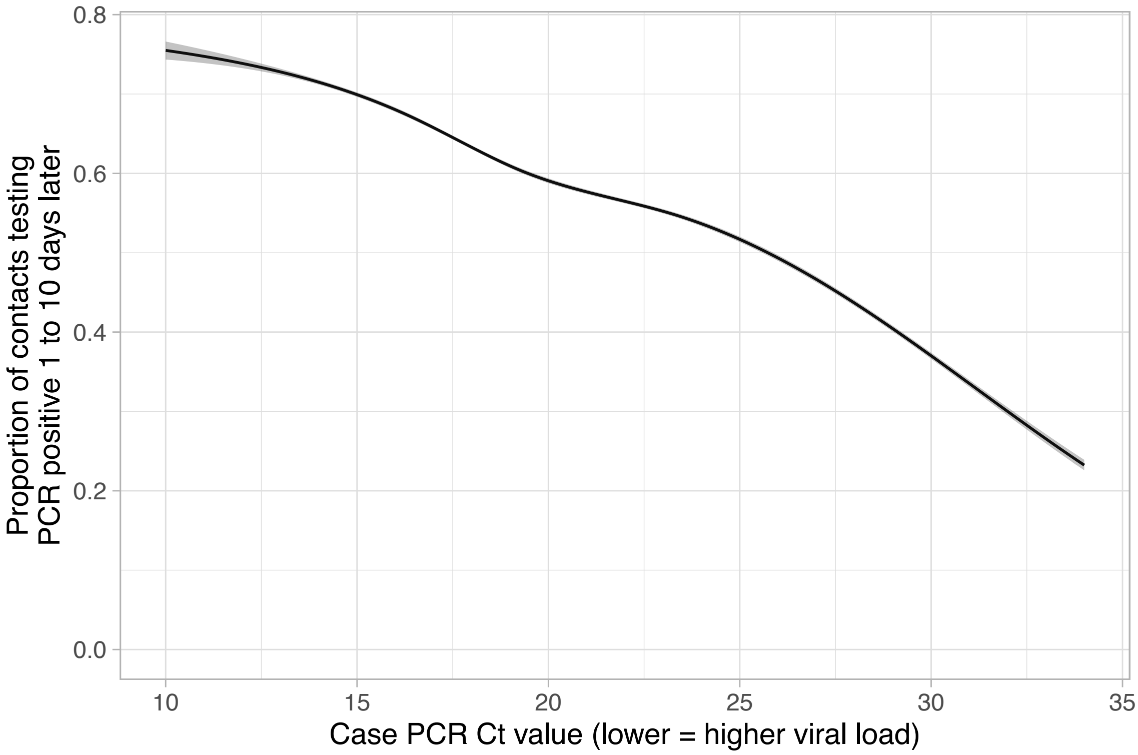


**Figure S14. Sensitivity analysis (restricting to contacts with a PCR test): univariable relationship between index case Ct value and the proportion of contacts testing PCR positive**. The shaded are shows the 95% confidence interval.

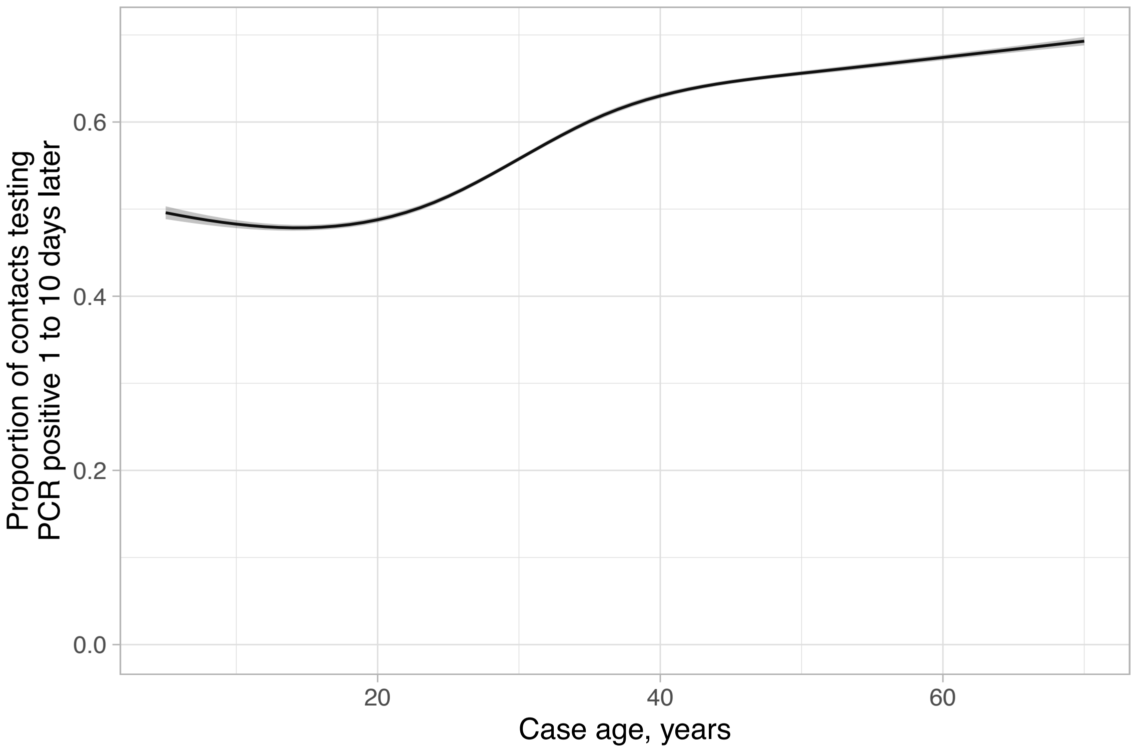


**Figure S15. Sensitivity analysis (restricting to contacts with a PCR test): univariable relationship between index case age and the proportion of contacts testing PCR positive**. The shaded are shows the 95% confidence interval.

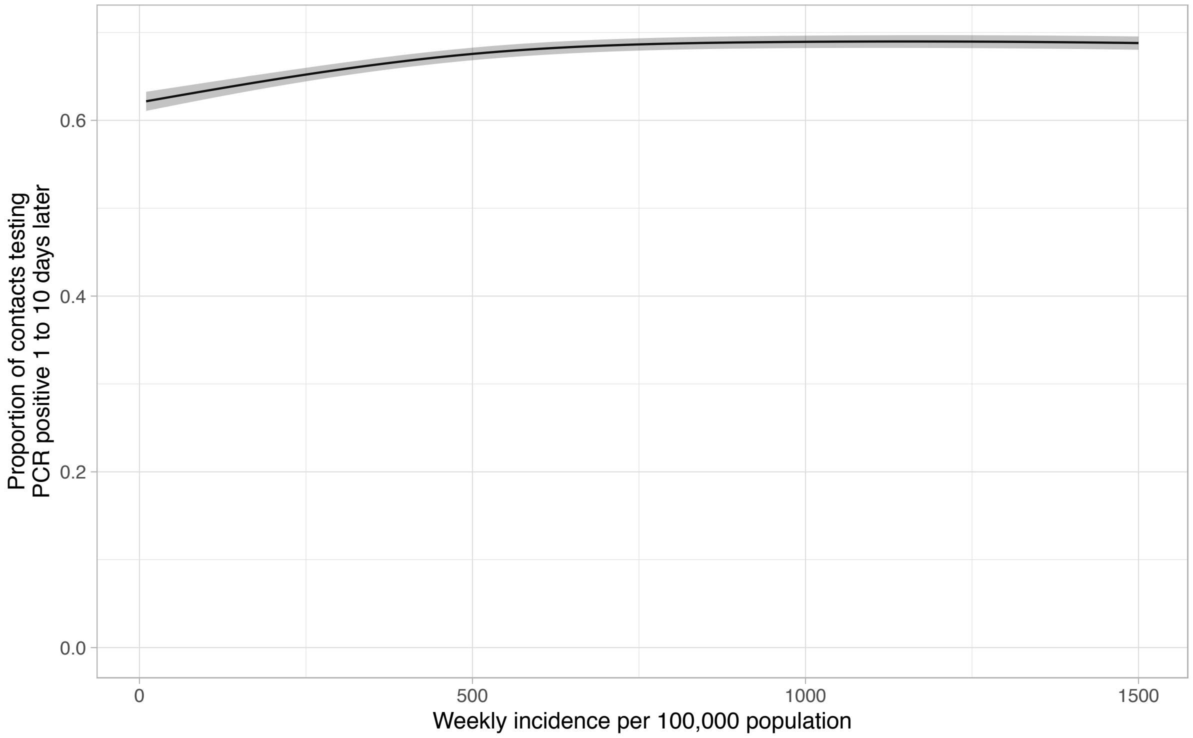


**Figure S16. Sensitivity analysis (restricting to contacts with a PCR test): multivariable relationship between weekly incidence at the contact's home address and the proportion of contacts testing PCR positive**. The shaded are shows the 95% confidence interval. Adjustment has been made for variables listed in Table S2.

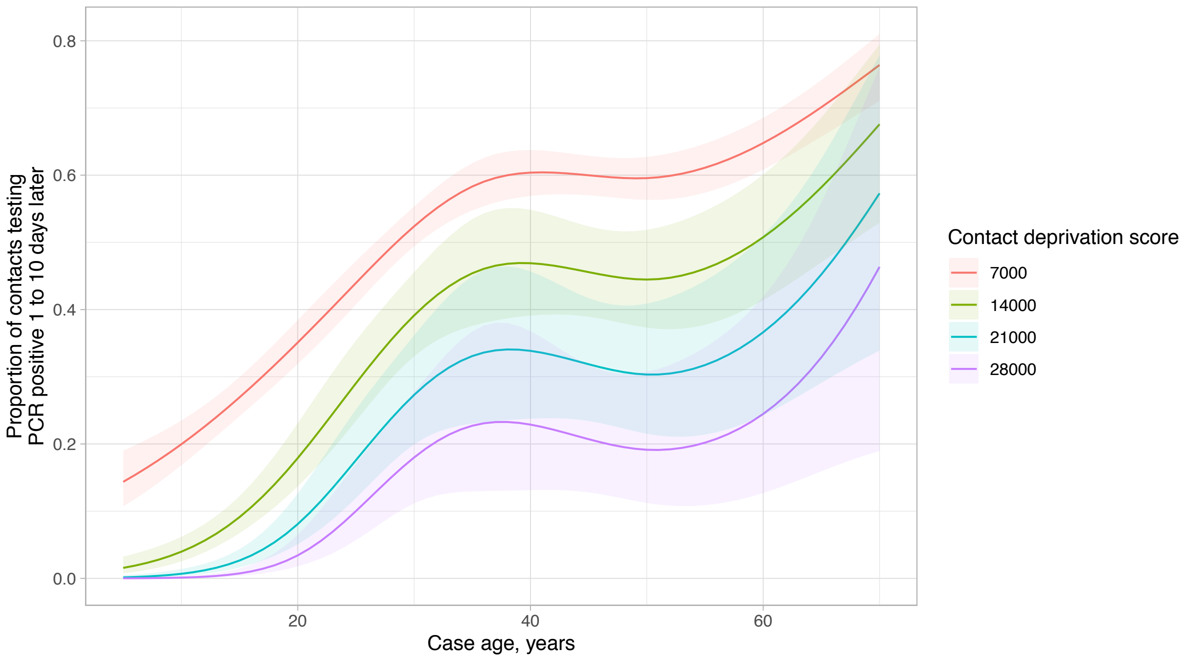


**Figure S17. Sensitivity analysis (restricting to contacts with a PCR test): Relationship between index case age and the proportion of contacts with a PCR positive result, by contact deprivation score**. Model predictions are adjusted for the factors in Table S2. The shaded area indicates the 95% confidence interval. Deprivation score is shown using the index of multiple deprivation rank, where 1 is the most deprived.

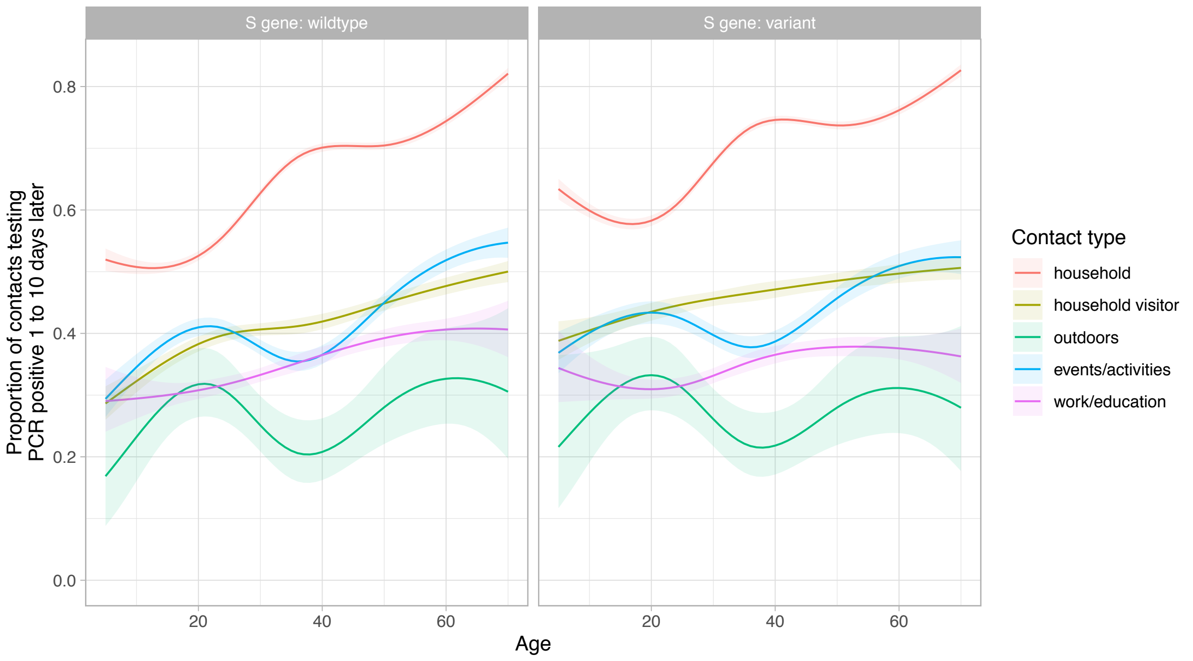


**Figure S18. Sensitivity analysis (restricting to contacts with a PCR test): Relationship between index case age and the proportion of their contacts with a PCR positive result, by contact type and S gene target failure**. Model predictions are adjusted for the factors in Table S2. The shaded area indicates the 95% confidence interval.

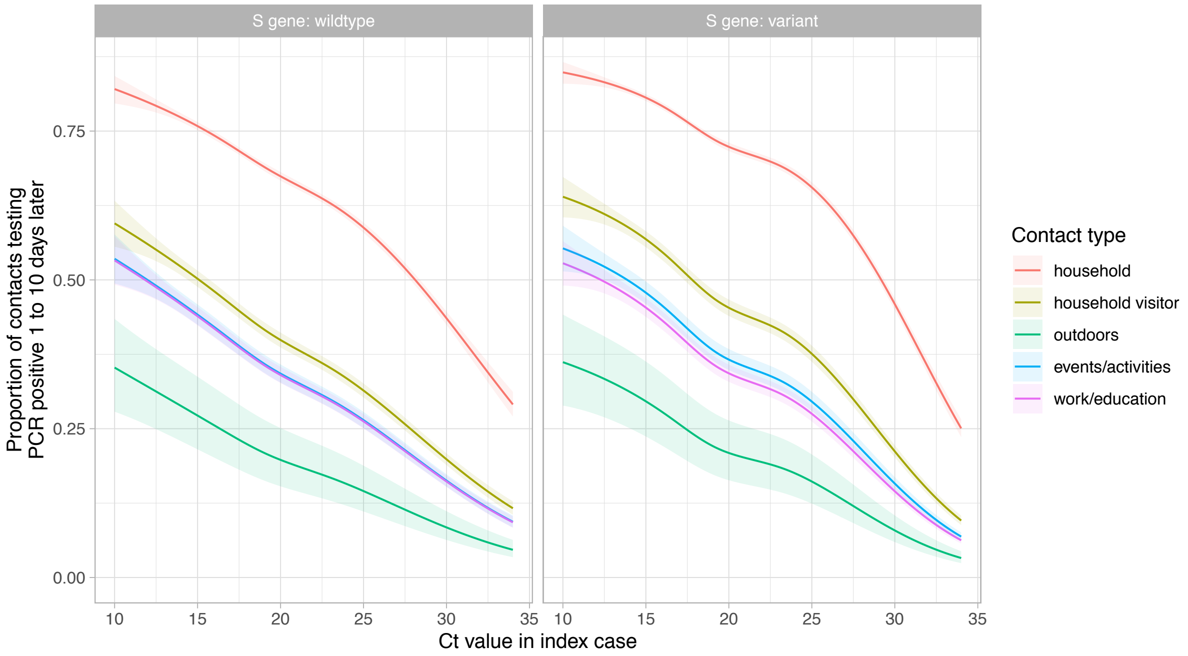


**Figure S19. Sensitivity analysis (restricting to contacts with a PCR test): Relationship between PCR cycle threshold (Ct) value in cases and the proportion of their contacts with a PCR positive result, by contact type and S gene target failure**. Model predictions are plotted after adjustment for index case age (set to the median value, 35 years), case ethnicity (set to white), index of multiple deprivation score at contact's home address (set to median, 14,465), incidence at contact's home address (set to median 350 cases per 100,000 population per week) and index case sex (set to female). The shaded area indicates the 95% confidence interval.

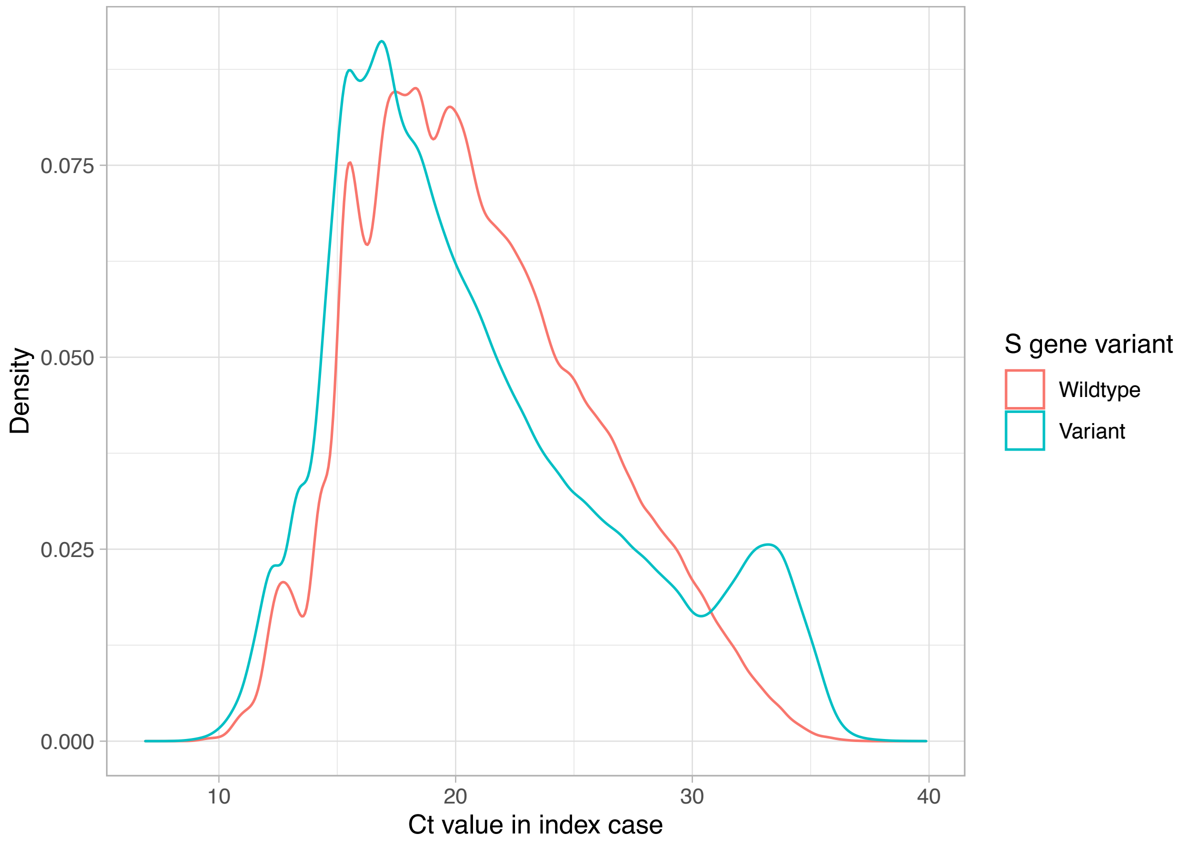


**Figure S20. Distribution of mean Ct values by SGTF**. Mean Ct values were calculated across all non-missing SARS-CoV-2 PCR targets.

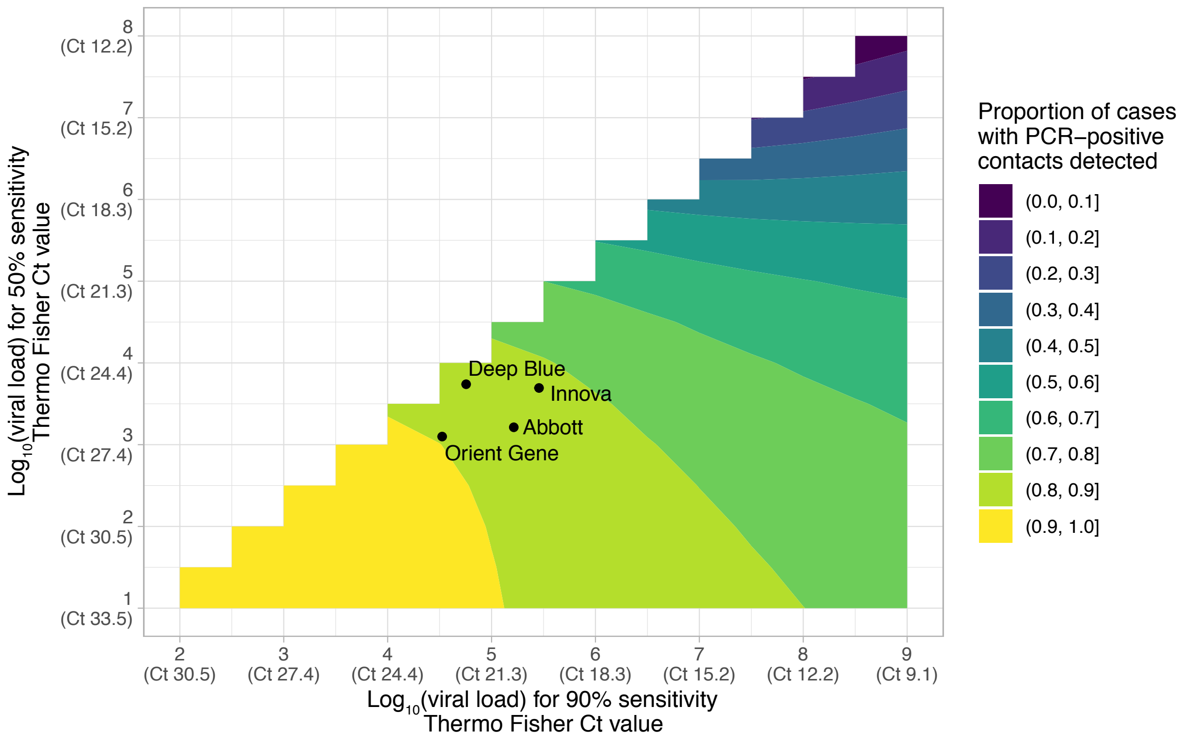


**Figure S21. Simulated performance for a range of LFDs.** The sensitivity of a LFD by viral load can be summarised using a logistic model with two parameters, a slope and intercept. The slope and intercept can be transformed into two alternative parameters, e.g. the viral load at which 50% of all individuals are LFD positive and the viral load at which 90% of all individuals are LFD positive. For a given combination of two parameters we simulate the proportion of cases with PCR-positive contacts that are detected by LFD, the mean result over 100 simulations is plotted. Thermo Fisher TaqPath assay equivalent Ct units are shown using the formula: $\log_{10}$(viral load) = 12.0 - 0.328*Ct. The performance of 4 LFDs (Figure S1, Figure 5) are overlaid on the simulation results.

### Supplementary tables

| Variable | Overall, n = 1,768,246[1] | Cases with ≥1 unique contact, n = 1,064,004[1] | No recorded contacts, n = 439,482[1] | Only shared contacts, n = 264,760[1] |
|---|---|---|---|---|
| Case sex | | | | |
| Female | 848,631 (48%) | 560,557 (53%) | 151,915 (35%) | 136,159 (51%) |
| Male | 763,001 (43%) | 476,967 (45%) | 162,674 (37%) | 123,360 (47%) |
| Not specified | 156,614 (8.9%) | 26,480 (2.5%) | 124,893 (28%) | 5,241 (2.0%) |
| Case ethnic group | | | | |
| Asian | 191,922 (11%) | 128,218 (12%) | 34,001 (7.7%) | 29,703 (11%) |
| Black | 42,604 (2.4%) | 27,658 (2.6%) | 10,194 (2.3%) | 4,752 (1.8%) |
| Mixed | 40,651 (2.3%) | 27,263 (2.6%) | 7,390 (1.7%) | 5,998 (2.3%) |
| Not available | 384,958 (22%) | 136,918 (13%) | 214,432 (49%) | 33,608 (13%) |
| Other | 23,866 (1.3%) | 15,682 (1.5%) | 4,981 (1.1%) | 3,203 (1.2%) |
| White | 1,084,245 (61%) | 728,265 (68%) | 168,484 (38%) | 187,496 (71%) |
| Case age | 36 (23 - 51) | 36 (24 - 51) | 35 (24 - 51) | 37 (22 - 53) |
| Unknown | 16 | 9 | 4 | 3 |
| Days from symptom onset to test in case | 2 (1 - 3) | 2 (1 - 3) | 2 (1 - 4) | 2 (1 - 3) |
| Unknown | 332,455 | 94,062 | 212,807 | 25,586 |

**Table S1. Demographics of cases with ≥1 unique contact included in the study, and groups excluded with no contacts or only shared contacts**. Cases without recorded contacts had higher amounts of missing data on sex and ethnicity, in part reflecting it was not possible to complete contact tracing for all cases. As such the proportion of cases with no contacts, as opposed to no recorded contacts, is likely to be lower than reported. Those with only shared contacts were broadly similar to those with ≥1 unique contact. [1]Frequency (%) or median (IQR)

| Variable | | Univariable | | | Multivariable | | |
|---|---|---|---|---|---|---|---|
| | | Odds ratio | 95% CI | p-value | Odds ratio | 95% CI | p-value |
| **Incidence contact's home address, per 100,000 population*** | 50 (baseline) | 1.00 | | <0.001 | 1.00 | | <0.001 |
| | 100 | 1.05 | 1.04 - 1.06 | | 1.16 | 1.14 - 1.17 | |
| | 200 | 1.14 | 1.13 - 1.14 | | 1.20 | 1.18 - 1.21 | |
| | 400 | 1.23 | 1.23 - 1.24 | | 1.23 | 1.22 - 1.25 | |
| | 600 | 1.27 | 1.26 - 1.28 | | 1.24 | 1.22 - 1.25 | |
| **Deprivation rank at contact's home address (lower = more deprived)** | per 10,000 increase | 0.88 | 0.87-0.90 | <0.001 | Interaction with age, see Figure S17 | | |
| **Case Ct value (lower = higher viral load)*** | 10 (baseline) | 1.00 | | <0.001 | Interaction with SGTF, interaction with age, see Figure S18 | | |
| | 15 | 0.93 | 0.92 - 0.93 | | | | |
| | 20 | 0.78 | 0.78 - 0.79 | | | | |
| | 25 | 0.69 | 0.68 - 0.69 | | | | |
| | 30 | 0.49 | 0.49 - 0.50 | | | | |
| **S gene target failure (SGTF)** | Wildtype (baseline) | 1.00 | | | Multiple interactions, see other rows | | |
| | S gene variant | 1.33 | 1.31 - 1.35 | <0.001 | | | |
| **Case sex** | Female | 1.00 | | | 1.00 | | |
| | Male | 1.10 | 1.09 - 1.11 | <0.001 | 1.00 | 0.99 - 1.01 | 0.95 |
| | Not specified | 1.09 | 1.04 – 1.14 | <0.001 | 1.07 | 1.02-1.13 | 0.01 |
| **Case age*** | 30 years (baseline) | 1.00 | | <0.001 | Interactions between SGTF and contact type, SGTF and age, contact type and age, see Figures S18, S19 | | |
| | 10 years | 0.87 | 0.86 - 0.87 | | | | |
| | 50 years | 1.18 | 1.17 - 1.18 | | | | |
| | 70 years | 1.24 | 1.23 - 1.25 | | | | |
| **Contact event** | Household (baseline) | 1.00 | | | | | |

| | | | | | | | |
|---|---|---|---|---|---|---|---|
| | Activities and events | 0.36 | 0.35 - 0.37 | <0.001 | | | |
| | Household visitor | 0.39 | 0.38 - 0.40 | <0.001 | | | |
| | Work or education | 0.28 | 0.27 - 0.29 | <0.001 | | | |
| | Outside | 0.19 | 0.17 - 0.21 | <0.001 | | | |
| **Case ethnicity** | White (baseline) | 1.00 | | | 1.00 | | |
| | Asian | 1.50 | 1.47 - 1.54 | <0.001 | 1.27 | 1.24 - 1.30 | <0.001 |
| | Black | 1.26 | 1.20 - 1.32 | <0.001 | 1.21 | 1.15 - 1.27 | <0.001 |
| | Mixed | 0.99 | 0.95 - 1.04 | 0.78 | 1.13 | 1.09 - 1.19 | <0.001 |
| | Other | 1.31 | 1.24 – 1.40 | <0.001 | 1.17 | 1.10 - 1.25 | <0.001 |
| | Not available | 1.17 | 1.15 – 1.29 | <0.001 | 1.10 | 1.07 - 1.13 | <0.001 |

**Table S2. Univariable and multivariable associations with the proportion of contacts who underwent PCR testing having a positive result**. *Incidence, case Ct value, and case age were included as non-linear terms with 4, 5 and 5 default spaced knots respectively (see Figures S13-S15 for univariable relationships). The multivariable relationship for incidence is plotted in Figure S16. CI, confidence interval.

| Lateral flow device | Time period | Proportion of cases identified by LFD | 95% Confidence interval |
|---|---|---|---|
| Deep Blue | September 2020 - November 2020 | 85.3% | 85.2 - 85.4% |
| Deep Blue | December 2020 - February 2021 | 86.3% | 86.2 - 86.4% |
| Innova | September 2020 - November 2020 | 82.2% | 82.0 - 82.3% |
| Innova | December 2020 - February 2021 | 83.5% | 83.3 - 83.7% |
| Orient Gene | September 2020 - November 2020 | 89.2% | 89.0 - 89.3% |
| Orient Gene | December 2020 - February 2021 | 89.8% | 89.6 - 89.9% |
| Abbott | September 2020 - November 2020 | 85.3% | 85.1 - 85.4% |
| Abbott | December 2020 - February 2021 | 86.2% | 86.1 - 86.4% |

**Table S3. Simulated lateral flow device (LFD) performance before and after the widespread emergence of the B.1.1.7 variant**.
